# An investigation of multisensory perception of surrounding space in aided congenitally hearing impaired

**DOI:** 10.1101/2024.06.02.24306672

**Authors:** Adi Snir, Katarzyna Cieśla, Rotem Vekslar, Amir Amedi

**Author notes:** equal contribution (Adi Snir, Katarzyna Cieśla).

## Abstract

We tested auditory spatial motion localisation in congenitally hearing impaired adult users of bilateral cochlear implants, and other hearing assistive devices. The group showed severely impaired capabilities despite extensive device use, emphasizing the role of *nature* in sensory development. We then investigate whether the deficit is maintained for other sensory modalities, by using an in-house sensory substitution device that provides weighted vibrotactile cues on fingertips to induce 3D spatial motion perception. The performance was significantly higher, both in the combined audio-tactile task and the tactile task itself, with accuracy comparable to typically hearing subjects. With touch, we also showed considerably fewer front-back and right-left confusions. The rapid audio-tactile binding and availability of 3D space representation through touch, point to the significant role of *nurture* in spatial perception development and its amodal nature. The findings show promise towards advancing multisensory solutions for spatial hearing rehabilitation.

**Highlights:**

– Auditory motion localisation is severely impaired in aided congenitally hearing impaired, supporting the role of *nature* towards spatial development;
– Binding auditory and tactile information enhances auditory spatial performance, supporting the role of *nurture*;
– Hearing impaired individuals perform 360° motion localisation through touch with accuracy similar to typically hearing;
– Surrounding spatial representation never before experienced in congenitally hearing impaired is rapidly available through an alternate modality

## Introduction

Imagine a world where you can perceive sounds, perhaps even recognize what they are, but you find it impossible to pinpoint where exactly they are coming from. When someone calls you from the left, you turn right; or even worse, a car approaches you from out of your sight, you hear the noise, but cannot use the basic instinct to avoid coming in contact with it. These types of everyday challenges are constantly faced by the hearing-impaired (HI) population, including experienced bilateral cochlear implants (BiCI) users (see e.g. Anderson et. al, 2022, Dorman et. al, 2016, Shafiro et. al, 2015, Zheng et. al, 2022, Moua et. al, 2019). When all sensory modalities are available, they all provide information about the 3D space, thus rendering spatial perception an amodal or multisensory experience (Spence & Di Stefano, 2024, Bruns & Röder, 2023). Audition, however, is the only sensory modality capable of perceiving external sources from all angles simultaneously (including behind the head) without moving the head or the body (Schnupp, Nelken & King, 2011). Meanwhile, vision is only frontally oriented and the tactile system (traditionally) only represents information within the peripersonal space, i.e. within reach. Does this entail that lack of optimal spatial auditory experience prevents proper development of 3D spatial representation?

According to some seminal works, sensory-specific information must be acquired early in life during “critical periods” in order for sensory functionalities to properly develop (Heimler & Amedi, 2020, Hubel, & Wiesel, 1962). Some authors also indicate that an early sensory exposure is necessary for multisensory integration to occur (Gori et. al, 2021, Bruns et. al, 2022; King, 2004). Findings in deaf or hearing impaired individuals, including those equipped with hearing devices (such as hearing aids/HAs or cochlear implants, CIs), who perform poorly in spatial localization tasks, even following multiple years of access to binaural information seem to confirm that assumption for the auditory system function. This also underscores the role of *nature* as opposed to *nurture* in sensory development (Kral & Sato, 2020). Most spatial localisation studies in the hearing impaired, however, use non-ecological static sounds positioned in the frontal field (e.g. Pavani et. al, 2017, Anderson et. al, 2022, Dorman et. al, 2016, Shafiro et. al, 2015, Zheng et. al, 2022), with only few recent ones taking the entire surrounding 360° space into account, including the area behind the head (e.g. Coudert et. al, 2022, Mueller et. al, 2014, Nisha, Uppunda & Kumar, 2023), as well as dynamic sound scenes (eg. Dwyer et. al, 2021, Fischer et. al, 2021, Moua et. al, 2019).

Auditory localization is based on binaural cues derived from constantly performed comparisons of the Interaural Time Differences (ITDs) and the Interaural Level Differences (ILDs) of the sources arriving at the two ears. Additional monaural cues are available due to the individual’s shape of the ear, head and torso (Middlebrooks, 2015). To calculate the auditory source position accurately, the auditory system must perform calculations at extremely high speeds, and even more so when sources are in motion (Schnupp, Nelken & King, 2011, Carlile & Leung, 2016).

Hearing impairments affect sound localisation by either limiting the perceived frequency ranges needed for localization or by distorting the incoming sounds, and thus also the binaural cues. In addition, auditory devices (HAs, CIs), optimized for speech comprehension, do not preserve the localization cues well, in part due to the specific sound encoding algorithms, signal compression, and gain adjustments (Johnson, Xu & Cox, 2017, Akeroyd, 2015, Zheng et. al, 2022). Furthermore, the post-treatment training of HI focuses mainly on speech comprehension (Carlyon & Goehring, 2021, Lu, Zhang, & Gao, 2019)

Given these limitations, can an alternative sensory modality be considered to represent spatial information in a manner similar to the auditory system? We turned to the animal kingdom for inspiration. In fact, there are animals, such as elephants, who can use seismic waves registered through their feet as tactile vibrations to detect distant sound sources. To enable source localization, an elephant can even lift one foot off the ground to effectively perform triangulation (e.g. O’Connell-Rodwell, 2007). Another interesting mechanism exists in some species of fish equipped with a lateral line of epithelial cells on the sides of the body which encode fluid motion in water.. It is speculated that the inner ear in mammals (and especially the sensory cells themselves) developed from a vestibular organ stemming from the lateral line (Kalmijn, 1989).

Furthermore, the auditory and the tactile systems in both mammals and humans alike, are densely connected at all levels of the central nervous system, and both are capable of encoding vibrations in a shared frequency range of 30-1000Hz through mechanoreceptors (e.g. Ro, Ellmore & Beauchamp, 2013, Li Hegner et. al, 2010, Kayser et. al, 2005, Beauchamp et. al, 2008, Caetano & Jousmäki, 2006, v. Békésy 1957) with a common ancestral lineage claimed between certain tactile mechanoreceptors and cochlear hair cells, Yu et. al, 2021).

To represent 3D motion specifically, we developed an in-house 3D technology with a touch-motion algorithm (TMA) that provides spatial cues through vibro-tactile stimulation on four fingertips of two hands. Similar to the auditory system, TMA performs vector-based level-weighting on multiple vibrotactile actuators to represent external sources. Since 3D localization via weighted cues is typically associated with hearing, our solution can be considered a *sensory substitution device* (SSD), i.e. a technology using an alternative sensory modality to convey information typically perceived by another natural sense (other examples include e.g. an electrotactile implant on the tongue for navigation in blind, or “soundscapes” representing visual objects (Bach-y-Rita et. al, 1969, Meijer, 1992, Kupers & Ptito, 2011). The research is also a continuation of the classical works from 1970/80s when simple set-ups for localisation using vibrotactile cue weighting and vibrations delivered on various body parts were developed, with promising findings (review in Borg, 1997, v. Békésy 1957).

Our prior work using TMA shows that typically hearing (TH) individuals are capable of using weighted tactile stimulation to represent spatial motion with accuracy comparable to that for the corresponding auditory task, along with enhanced performance for sensory integration in noise (Snir et. al, 2024). In the current study, we expand our investigation into HI individuals. Specifically, our experiment measures localization capabilities of auditory, tactile and paired auditory-tactile sources moving on a 360° azimuthal plane in a group of BiCI users with congenital hearing loss and early fitting of one CI. In addition, we test single subjects with one CI, a bimodal CI-HA solution and bilateral HAs. We suggest this new approach, based on studies using SSDs in congenitally blind individuals which showed that certain *visual* skills are learnable in adulthood following hours of training (indicating the role of *nurture vs nature* in the development of sensory functionalities despite lack of early experience; e.g. Heimler & Amedi, 2020).

Based on the existing research in this population, we hypothesize that the CI/HA users will display compromised auditory localization abilities, as compared to those with typical hearing (TH). At the same time, we speculate that their performance will improve when auditory and tactile cues are both present (as compared to audio only). This would show that a new and untrained sensory input can be integrated with one that has been used throughout lifetime and in evolution. Such a prediction is also viable in consideration of the “rule of inverse effectiveness” of multisensory integration, which posits that the benefit of providing information through an additional sensory modality is proportional to the reliability of the original sense (Meredith & Stein, 1983, Holmes, 2007). An effect which was also found in the typically hearing population in our prior work when noise was added to the audio environment during an audio-tactile 3D spatial task (Snir et. al, 2024). For tactile spatial localisation in the congenitally hearing impaired group, we posit that if an early experience of 3D space through hearing is necessary for the development of 3D spatial perception in general, then performance in the tactile task will also be compromised. At the same time, if the tactile modality is capable of performing the new 3D task, this will strengthen the role of *nurture* in building a 3D space representation and emphasize its amodal character.

## Results

### Performance in the spatial motion localisation tasks

In the spatial localisation tasks, the participants were asked to localize sounds moving on a 360° azimuth.

#### Hearing impaired individuals perform significantly worse than the typically hearing controls in auditory spatial motion localization

##### Scores in A1 (baseline) condition

BiCI users (N=10) had the following scores in the Audio 1 (baseline) condition: A1 mean score (0.58+/− 0.018), A1 median (0.62, IQR=0.50). When analyzed jointly, the CI/HA users (N=19) had very similar scores; A1 mean score (0.58+/−0.19), A1 median (0.57, IQR=0.38). TH individuals (N=29) had the following results in the same task: A1 mean score (0.90+/−0.19), A1 median (1, IQR=0.25). The BiCI users showed significantly lower auditory scores, as compared to the TH in A1 (z=16.8, p<0.001; large effect size, Cohen’s d=1.3, power 99%), and the same was found for all CI/HA users jointly (z=19.9, p<0.001). All the results were significantly above chance (p<0.001). Figure 1 depicts the results.

**Figure 1.**
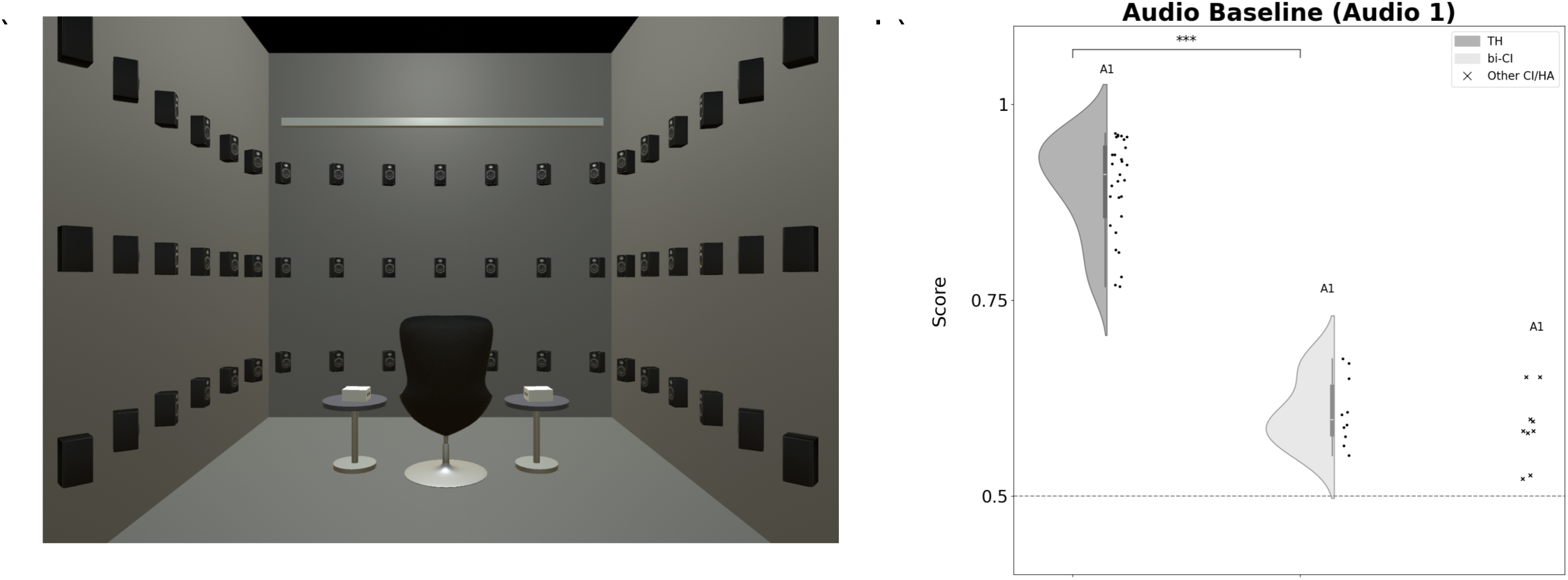
Auditory 3D motion localization: condition A1. a) Experimental set-up with 97 speakers on the walls & on the ceiling; b) Group scores in bilateral cochlear implant users (BiCI) in comparison to the typically hearing individuals; scores of other hearing device users (other CI/HA) represented as individual crosses on the right side. Statistical comparisons with a Wilcoxon signed-ranks test (*** p<0.001, ** p<0.01, *p<0.05). Data are represented as mean +/− SD.

##### Scores in A2 condition

In the A2 task, BiCI users had the following scores: A2 mean (0.55+/−0.18), A2 median (0.62, IQR=0.59). The scores of all CI/HA together were similar: A2 mean (0.59+/−0.012), A2 median (0.62, IQR=0.50). For the typically hearing individualis, the scores were: A2 mean (0.91+/−0.19), A2 median (1, IQR=0). BiCI A2 results were significantly lower than those found in TH (z=18.12, p<0.01; Cohen’s d = 1.3, power=99%; Mann Whitney tests), and the same was found for all CI/HA users jointly (z=20.1, p<0.001): All the results were significantly above chance (p<0.001).

No statistically significant differences were found between the A1 and A2 scores in either of the groups.

#### Hearing impaired individuals show significantly higher results when performing the task through touch, as compared to audio only

In the BiCI users, the T scores were: mean (0.78+/−0.16), median (0.875, IQR=0.25), and were significantly higher than the A1 results in this group (z=-7.35, p<0.001, Wilcoxon signed-ranks test). When analyzed together, the group of all CI/HA users (N=19) obtained the following very similar results in the T condition: mean score (0.79+/−0.011), median (0.87, IQR=0.25), and these were also significantly higher than the A1 results (z=11.4, p<0.001; large effect size, Cohen’s d=0.85, power = 98%). Within the TH group, the T results were: T mean (0.86+/−0.01), median (1, IQR=0.25) and were significantly lower than their A1 results (z=5.3; P<0.001; Wilcoxon signed-rank test). When the groups were compared with one another, Mann Whitney tests showed significantly lower scores in the participants with hearing loss, as compared to the TH subjects in T, both for BiCI users (z=4.11, p<0.001) and all the hearing impaired subjects jointly (z=4.15, p<0.001). All the results were significantly above chance (p<0.001). See Fig 2.

**Figure 2.**
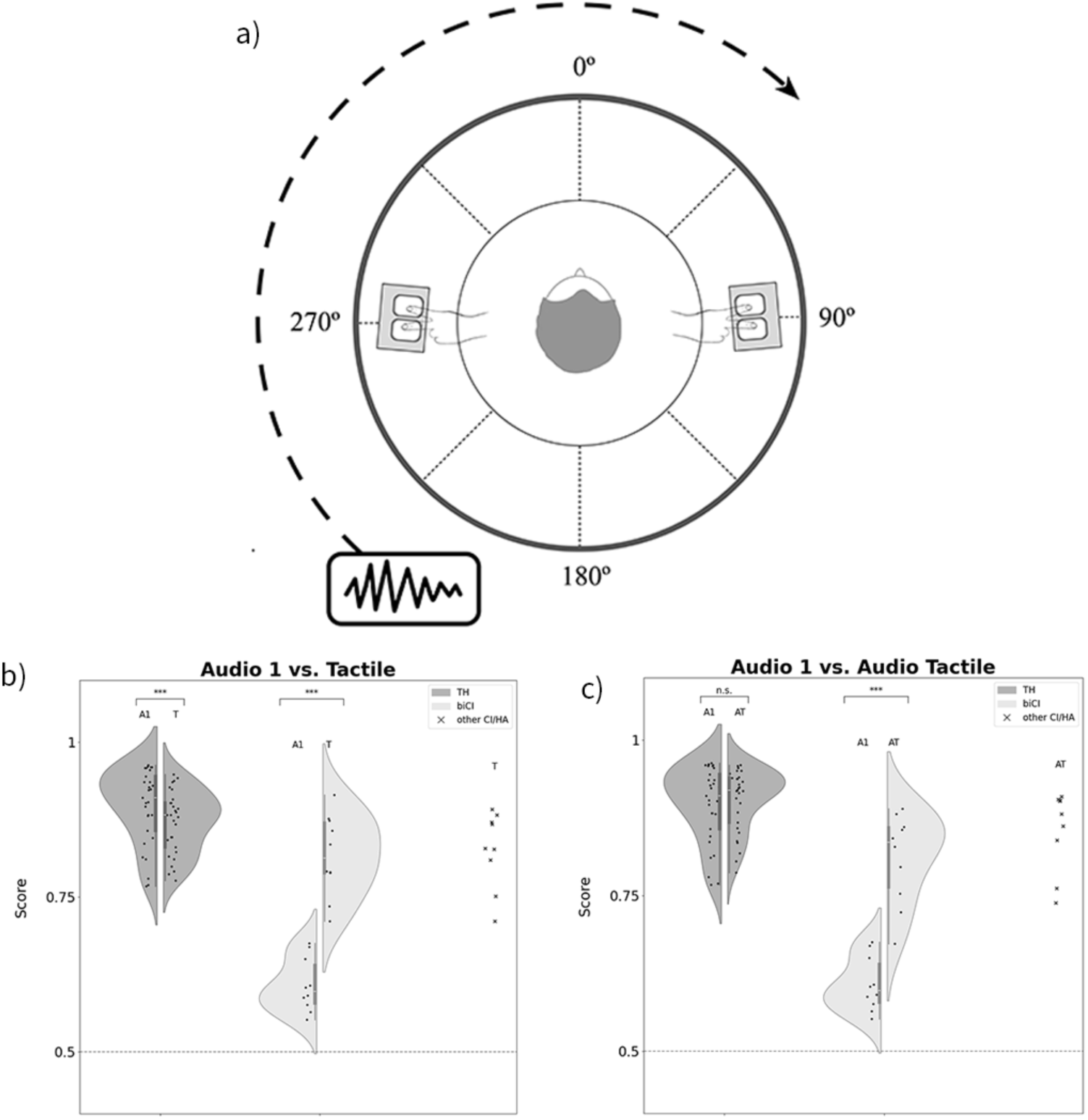
Auditory (A1), Tactile (T) and Audio-Tactile (AT) 3D motion localization. a) Experimental set-up with fingertips inserted in the two vibrotactile devices on the sides of the participant, and an example trajectory of a moving stimulus; b) and c) Group scores in the hearing impaired individuals: bilateral cochlear implant users (biCI) and other hearing impaired (CI/HA), and the typically hearing subjects. Statistical comparisons with a Wilcoxon signed-ranks test (*** p<0.001, ** p<0.01, *p<0.05). Data are represented as mean +/− SD.

#### Hearing impaired individuals show significantly higher results when performing a multisensory task, as compared to audio only

During the audio-tactile test condition, bilateral cochlear implant users had the following results: mean score 0.79+/−0.16; median 0.87, IQR=0.25). The scores were significantly higher than the baseline A1 scores (Z=8.27, p<0.001), indicating multisensory enhancement. The results of the entire hearing impaired group were almost the same (mean score 0.81+/−0.011; median 0.87, IQR=0.25) and also significantly higher than their baseline A1 scores (z=13.15, p<0.001). The group of TH scored: AT mean (0.93+/−0.006), median (1, IQR=0.25) and their AT results were not different from their baseline A1 scores (no multisensory enhancement). Mann Whitney tests showed significantly lower scores in all participants with hearing loss, as compared to the TH subjects in AT (z=9.1, p<0.001). And the same was shown for the group of BiCI users (z=9.1, p<0.001). All the results were significantly above chance (p<0.001). See Figure 2.

#### All hearing impaired individuals show a very similar pattern in their results

Figure 3 represents the results of all individual hearing impaired subjects.

**Figure 3.**
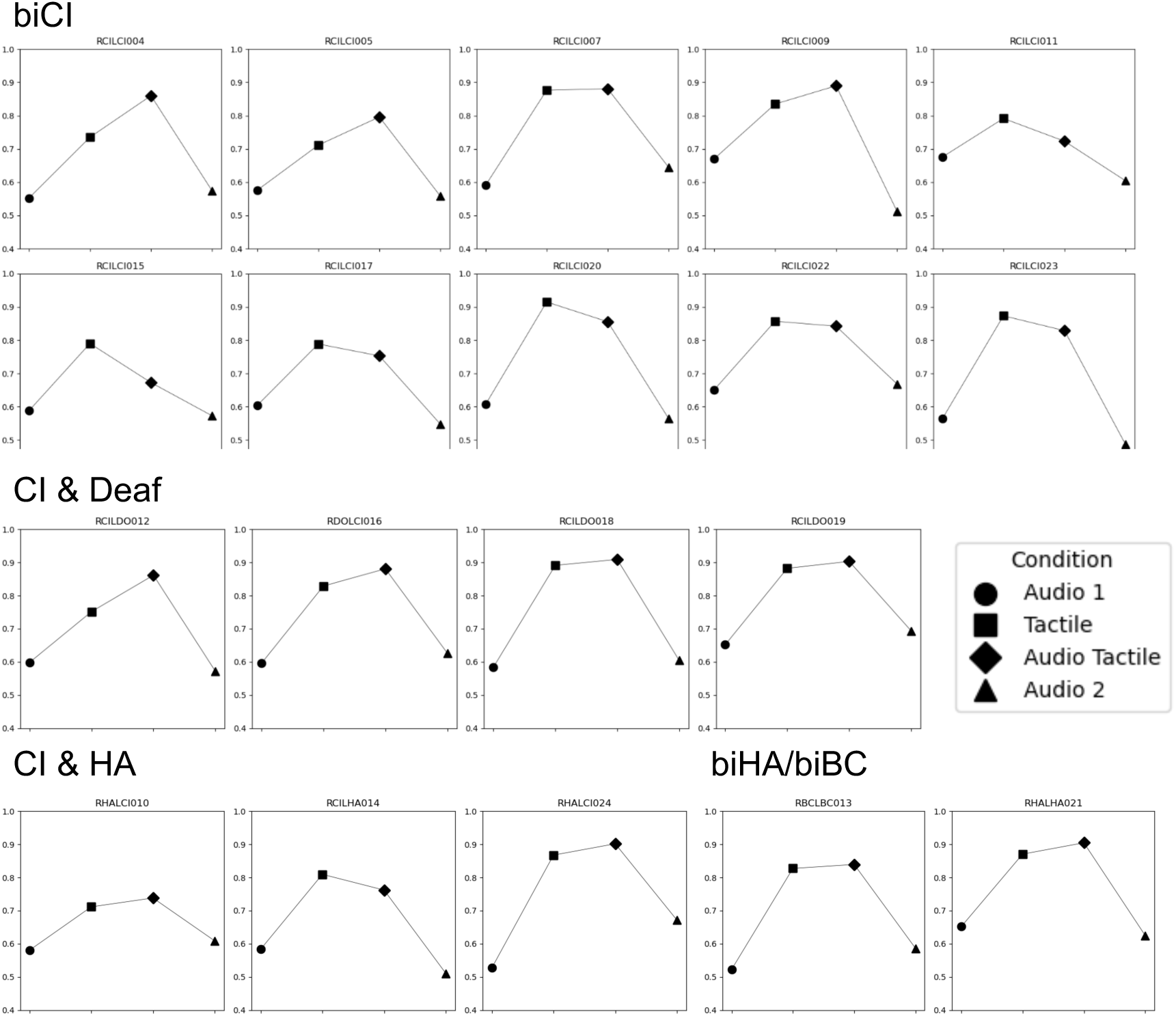
Single subject results in the spatial localisation task, showing performance in each test condition; CI – cochlear implant, HA – hearing aid, BC – bone conduction aid (rooftops)

#### Bilateral cochlear implant users make more FB/BF & RL/LR confusions, as compared to the typically hearing and when no tactile inputs are available

In BiCI users (N=10) the mean percentage of mistakes was as follows: a) in the A1 condition, right-to-left (RL) 15.4%+/−12.3, left-to-right (LR) 43.1%+/−30.2, front-to-back (FB) 73.1%+/−11.5, back-to-front (BF) 29.2%+/−13; b) in the T condition, RL 4.3%+/−7.3, LR 18%+/−18.5, FB 30.1%+/−29.1, BF 15.8%+-/-16.6, c) in the AT condition, RL 8.9%+-/11.5, LR 15.9%+/11.5, FB 16.1%+/−14.7, BF 11.6%+/−13.8, d) in the A2 condition, RL 19.3%+/−20, LR 51%+/−22.4, FB 71.6%+/−13.8, BF 36.3%+/−24.4.

In all CI/HA users all together (N=19) the mean percentage of mistakes was as follows: a) in the A1 condition, RL 21.48%+/−16.58, LR 44.9%+/−28.14, FB 69.2%+/−21.29, BF 33.8%+/−20.45; b) in the T condition, RL 6.5%+/−8.47, LR 18%+/−15.68, FB 31.8%+/−31.14, BF 12.7%+-/-13.62, c) in the AT condition, RL 6.45%+-/9.64, LR 16.22%+/13.26, FB 16.17%+/−18.6, BF 8.4%+/−10.87, d) in the A2 condition, RL 19.08%+/−16.14, LR 37.12%+/−25.12, FB 65.03%+/−23.06, BF 34.15%+/−30.29.

In TH individuals (N=29) the mean percentage of mistakes was as follows: a) in the A1 condition, RL 1.66%+/−3.7, LR 1.17%+/−3, FB 12.82%+/−22.3, BF 8.24%+/−14.89; b) in the T condition, RL 3.58%+/− 5.37, LR 3.11%+/−5.68, FB 6.24%+/−12.63, BF 13.02%+-/-13.9, c) in the AT condition, RL 1.63%+-/4.4, LR 2.78%+/6.37, FB 4.9%+/−8.8, BF 5.8%+/−12.77, d) in the A2 condition, RL 1.59%+/−4.2, LR 1.9%+/− 5.18, FB 12.34%+/−21.26, BF 8.09%+/−16.59.

Figure 4 represents the results of front-back and right-left confusions in TH and biCI participants.

**Figure 4.**
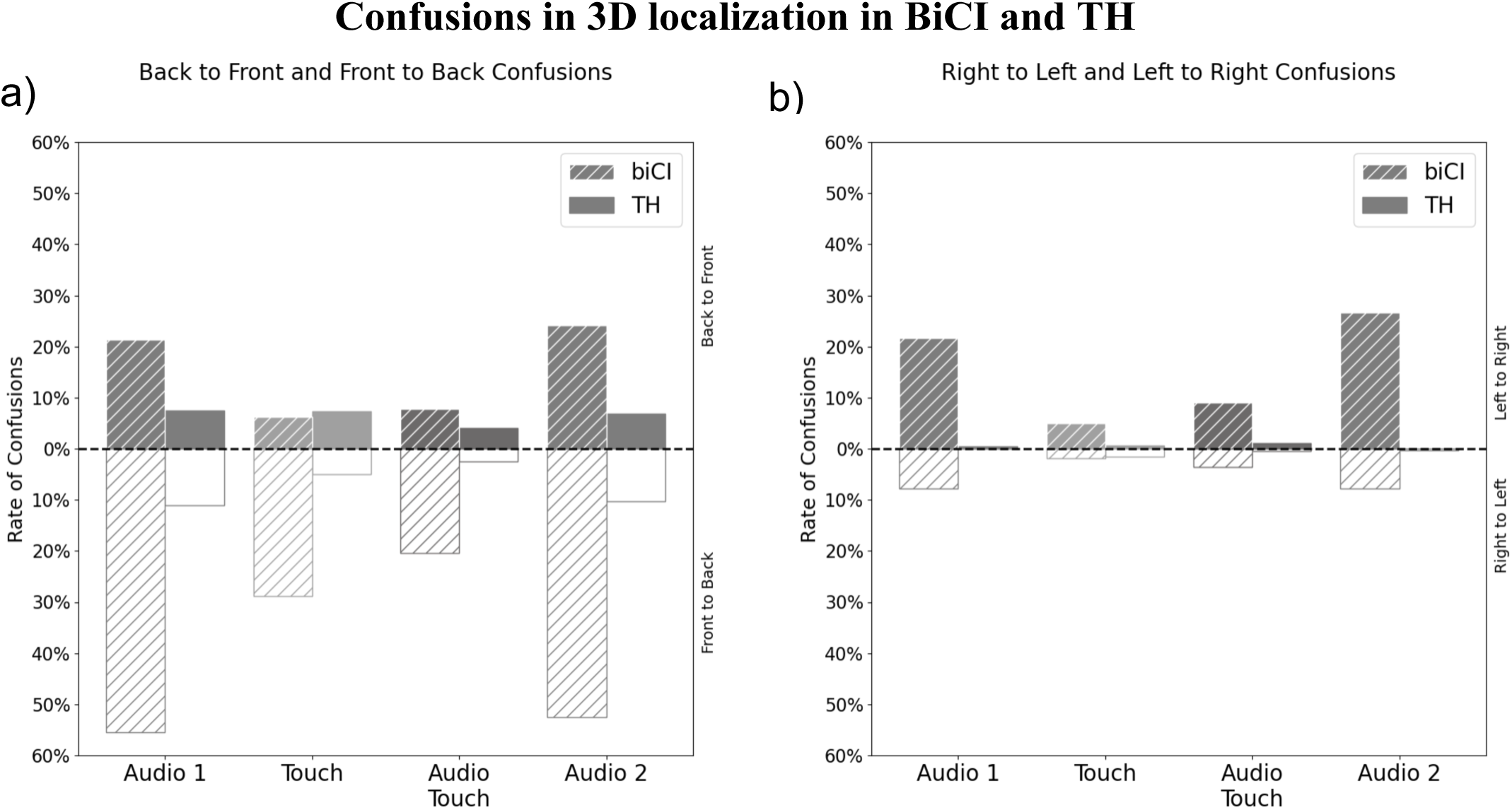
Front-Back and Right-Left confusions as percentage, in the typically hearing and in users of bilateral cochlear implants.

### Subjective questionnaire

The subjective questionnaire following participation in the task revealed some of the participants’ experiences. Questions and responses are summarized in Table 1.

**Table 1.**
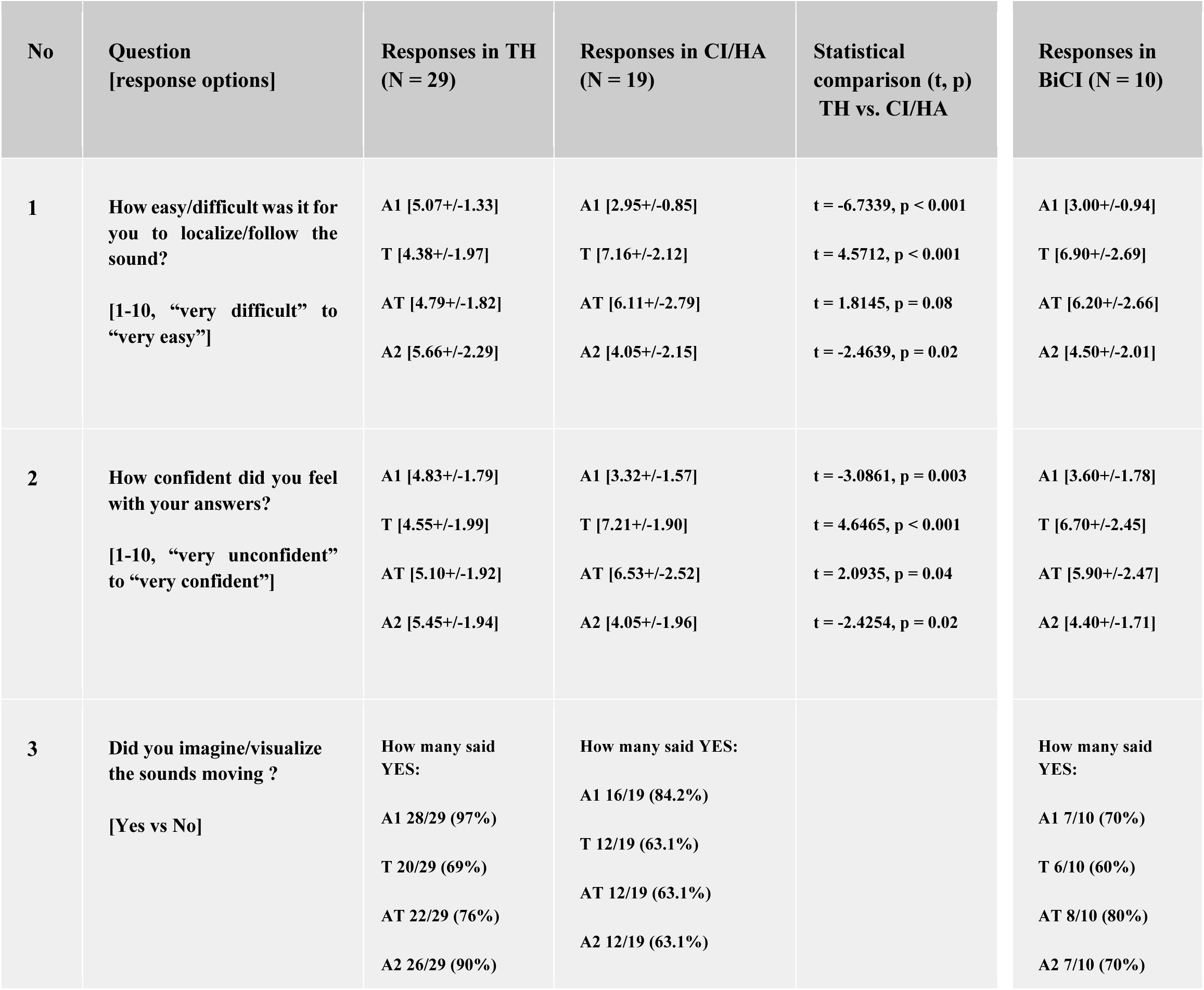
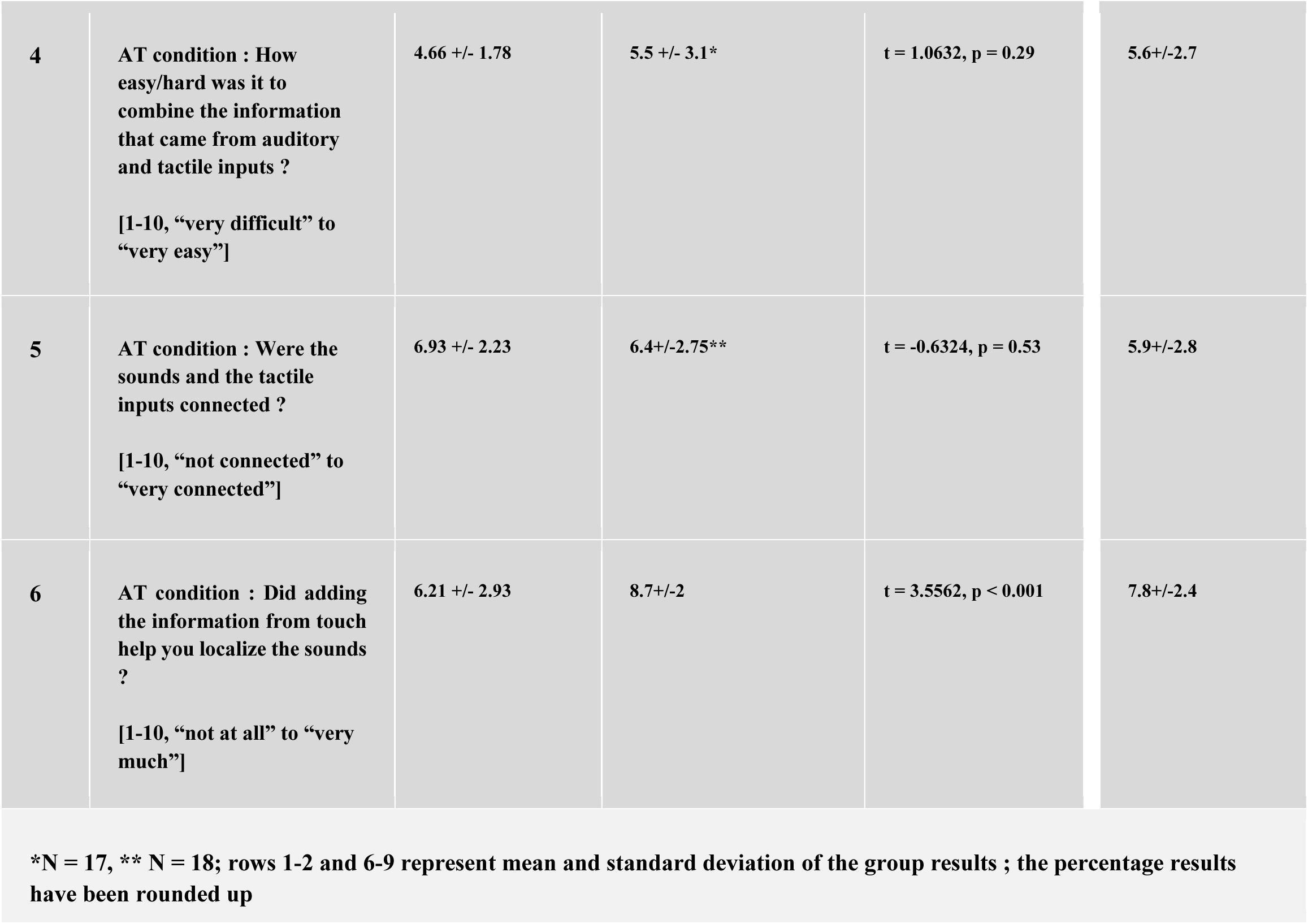
Subjective questionnaire and responses regarding test conditions in the motion localisation task.

### Correlation analysis

In the bilateral CI group (N=10) there was a negative correlation found between the age at fitting of both devices and the results in A1 test condition of the motion localization task (for the first CI: Kendall’s r=-0.53, p=0.0.019; for the second CI: Kendall’s r = –0.56, p=0.009). This result shows that an early implantation, and especially of the second implant can positively impact auditory spatial localisation.

### Additional psychological questionnaires

#### CTT

In the hearing impaired subjects (3 males, 13 females, age 29.13+/−11.5) the mean time of performing the CCT2 task was 71.06+/−19.35 seconds, and no misses or corrections were recorded. In the TH subjects (7 males, 19 females, age 23.37+/−1.4) the mean time for the same task was 74.73 +/− 16.73 seconds, and one participant made one mistake that he immediately fixed. These screening results indicate similar levels of performance in both groups and no issues with sustained or divided attention in neither group (D’Elia et. al, 1996).

#### NCIQ

The mean group NCIQ results of the 17 cochlear implant users were as follows: Basic Sound Perception (BSP; 60.58+/−17.75), Advanced Sound Perception (ASP; 77.36=5 +/− 13.7), Speech Production (SP; 63.23+/−13.57), Self-Esteem (SE; 69.85+/−11.54), Activity (A; 71.03+/−15.03), Social Interactions (SI; 70.3/-9.1). When analyzed separately, users of two cochlear implants (N=10) had very similar results: Basic Sound Perception (62.25+/−18.1), Advanced Sound Perception (79.75 +/− 13.61), Speech Production (67+/− 12.84), Self-Esteem (69.5+/−13.16), Activity (70+/−17.91), Social Interactions (73.25+/−9.72). A similar range of results was also found among the 7 users of one CI (and the other ear either with a HA or deaf). There were no consistent differences found in the scores between bilateral and unilateral CI users, and other participants, but the group sizes were small, preventing reliable statistical comparisons. The reported scores are similar to those shown by other research groups for CI users (Hinderink, Krabbe, & Van Den Broek, 2000, Santos, Couto & Martinho-Carvalho, 2017, Rasmussen et. al, 2022).

#### APHAB

Table S1 shows the results of the APHAB questionnaire of five single subjects using either two hearing aids or one hearing aid and one CI. The scores indicate that the participants varied significantly with respect to the perceived hearing-related handicap, both with and without the HA on. However, they all showed a benefit of using a HA in terms of Ease of Communication (i.e. communicating at home, in a public space), as well as in acoustic situations with Reverbnation (e.g. in a classroom or a theater; except for RHALCI024) or Background Noise (e.g. when there are multiple talkers). In the scale Aversiveness, they all reported that the handicap was higher with the HA turned on. This scale refers to situations such as traffic noise, sudden alarm sound, construction noise, etc. Most probably, these sounds are not available to the participants without a hearing aid.

## Discussion

### Auditory spatial capabilities are severely impaired in BiCI users and HI subjects

Our results are in agreement with and further contribute to the existing research on sound localization capabilities in BiCI users and other hearing impaired individuals (Pavani et. al, 2017, Anderson et. al, 2022, Dorman et. al, 2016, Shafiro et. al, 2015). Even though some prior research maintains that people with BiCIs can perform basic localization tasks better than those with unilateral CIs, here we show that this population still has impaired spatial capabilities and specifically when encountered with a real-world task involving surrounding sounds in motion (Zheng et. al, 2022, Andersson, 2015, Ludwig et. al, 2021). BiCI users perform the auditory task only slightly above chance (at approximately 60%; the large effect size also indicates that 92% of the HI group in general had lower scores than the TH subjects). Prior research on motion understanding in this population is scarce, but shows for example that it can be more challenging than localizing static sources (Moua et. al, 2019, Lundbeck et. al, 2017). Regarding unilateral CI users and bilateral HA users, we show similar levels of auditory performance in our complex setting (cf. a similar observation for localizing static sources in Dorman et. al, 2016).

The fact that auditory localization remains impaired despite years of auditory experience points towards the importance of *nature* for proper establishment of auditory spatial representation. This may be a result of only partial access to sound level cues (ILDs) and the severely obstructed temporal cues (ITDs; eg. Laback, Egger & Majdak, 2015, Kan, & Litovsky, 2015, Warnecke, Peng & Litovsky, 2020), both essential for successful sound localization. The role of *nature* is further emphasized in the correlation between age of fitting of the second CI, in particular, and auditory scores in our group of BiCI users (cf Schäfer et. al, 2021). With both CIs implanted early in life, a binaural model can be established and further refined with experience. While when the second CI is fitted later in life (in most participants here when they were teenagers), the spatial maps develop in a suboptimal manner. Indeed, early auditory deprivation in one ear, and prolonged stimulation in the other ear, has been shown to result in maladaptive aural preferences (e.g., Kral & Sato, 2013, Gordon, Wong & Papsin, 2013). Further investigation into the BiCI population is nevertheless needed in order to assess the potential of dedicated interventions for auditory recalibration which would prove effective towards development of auditory spatial abilities later in life (see similar discussion regarding blind in e.g. Gori, 2015; Bruns & Röder, 2023).

### Newly acquired tactile ability for spatial localization

While the auditory localization of 360° spatial motion was impaired among HI individuals (both biCIs and others), they achieved tactile accuracy nearly as high as the TH group (82% vs 84%), which was also close to the TH auditory performance (90%). The ability to perform a spatial localization task with such high accuracy through an alternative sensory modality, learned in adulthood, points to the role of *nurture* in the development of spatial representation.

This result also aligns with the broader framework of sensory substitution devices (SSD) and training programs which enable establishment of a novel connection between a computation and an atypical sensory modality (Bach-y-Rita et. al, 1969, Meijer, 1992, Kupers & Ptito, 2011, Maidenbaum et. al, 2014, Abboud et. al, 2014); and further indicate the brain’s ability to use novel information that was not introduced during the critical periods of development nor in evolution (Heimler & Amedi, 2020). Such rapid development of a new skill using another sense, with accuracy comparable to the life-long developed auditory modality is however rare, and a possible indication of spatial perception being amodal and highly malleable in nature (Spence & Di Stefano, 2024, Bruns & Röder, 2023, Motion localization in 360° has also hardly been investigated and even less so in the cochlear implant/hearing aid users.). This is in line with neuroimaging literature that shows shared neuronal mechanisms in the parietal cortex for spatial attention and in MT/V5 for motion processing, regardless of the applied sensory modality (Rezk et. al, 2020, Battal, 2018).

In addition, high tactile performance in the hearing impaired individuals demonstrates that an early sensory deprivation may not necessarily hinder the development of spatial representation, and further appropriate intervention may enhance such skills. Similarly, studies in congenitally blind individuals show effects of SSD-mediated training leading to acquisition of typically “visual” functions such as, e.g. navigation through vibrotactile inputs from an electronic walking cane (Maidenbaum et. al, 2014) or face recognition through specifically developed soundscapes (Abboud et. al, 2014, Arbel, Heimler, & Amedi, 2022). However, research using SSDs in people with partially impaired senses using assistive technologies, as in the current study, is significantly scarcer.

### Multisensory effects in HI in the spatial localization task

Our results show that spatial localization in the HI improves significantly when specifically developed tactile stimulation is provided in tandem with sounds (task performance at approximately 80% vs 60%, for AT and A1 in the both BiCI users and all hearing impaired subjects, respectively). We hypothesize that aligning the two sensory inputs in terms of content, spatial movement, and timing played a significant role in producing this multisensory effect (see the temporal and spatial principles of multisensory integration as outlined by Spence, 2010). The results also comply with the inverse effectiveness principle of multisensory integration (Meredith & Stein, 1983). The same effect can be seen in our prior work in typically hearing individuals when background noise was added to the task (Snir et. al, 2024).

The reported multisensory enhancement might also point to the role of early sensory experience and duration of device use in the development of multisensory integration. Other existing findings in CI users are mixed in that matter. In unilateral CI users, some works show no benefit of adding visual cues to an auditory vertical localization task (Pavani et. al, 2017), as well as impaired audio-tactile integration (parchment skin illusion) in cases of prolonged deafness (although accuracy improves with longer device use; Landry, Guillemot & Champoux, 2013, Guillemot & Champoux, 2014). Others indicate typical visual attention capture in BiCI users (Valzolgher et. al, 2023, Kamke et. al, 2014) and efficient integration of simple audio-tactile stimuli in both congenitally and late unilateral/bilateral CI recipients, leading to faster reaction times (Nava et. al, 2014). In the current experiment all participants were fitted with their first hearing device (either a CI or a hearing aid) in the first few years of life, and used it for many years. We speculate that access to sounds from early development, as well as intact visual and tactile sensory modalities allowed for the development of the observed multisensory integration (Bruns & Röder, 2023, King, 2004). That considered, the BiCI case is one in which participants used two technologically mediated sensory modalities (CIs with TMA) and were nevertheless able to integrate information quite seamlessly. Further investigation is required with other types of multisensory tasks, however.

Finally, while the auditory tasks were, as expected, reported in the subjective questionnaire as much more difficult for the HI group, the tactile and the audio-tactile spatial tasks felt significantly easier. The HI also found the tactile inputs much more helpful in localizing sounds than the TH control group. Indeed, compensating for missing auditory information by using tactile cues may be more familiar to HI individuals, such as for example when touching speaker membranes during listening to music (Tranchant et. al, 2017).

### Front-back and right-left confusions ameliorated by adding spatialized tactile inputs

One of the most frequently reported challenges with spatial localisation, both in the TH and the HI, are front-back confusions. The reason for this being that two sound sources originating from the same direction along the midline yield identical ILD and ITD comparisons (Schnupp, Nelken & King, 2011). In users of assistive devices they also stem from the spectral compression applied to the incoming signal and/or the placement of the microphones (Mueller et. al, 2014, Johnson, Xu & Cox, 2017). This is solved in real life by either taking advantage of the spectral cues, by moving the head so that the signals entering the ears differ or by turning to look at the sound source (McLachlan et. al, 2021, Yost, Zhong & Najam, 2015, Pastore et. al, 2018). In the current experiment, front-back (FB) and back-front (BF) confusions decreased significantly with tactile inputs complementing the auditory sources. In BiCI users, with a notably higher amount of errors than the TH, participants most frequently misreported the sources as coming from the back rather than the front. A likely reason being the lack of matching visual input for the frontal sources, a cue which this population is quite dependent on for localization (Bavelier, Dye & Hauser, 2006; Aurelie 2022). As for confusing the right with the left side, almost no such mistakes were made by TH subjects, with the lateral ITD and ILD cues being extremely dependable for these orientations (Middlebrooks, 2015). In some CI/HA users, however, there was a visible tendency to perceive some left oriented stimuli as coming from the right (17/19 HI participants preferred their right ear, as reported in the interview). The lateral confusions were also significantly ameliorated with the use of concurrent touch. Individual spatial orientation maps revealed similar tendencies in spatial perception of some of the HI, such as e.g. 8/10 BiCI and 2/3 CI/HA users (in sum 55%) making mistakes mostly when localizing auditory sources in the front or the front-left field (see Figure S1).

### Similarities between auditory and tactile processing and subjective subjective experiences

The specific tactile weighting and vibrations used in our study may have been critical for obtaining the observed high results. Prior research using more basic set-ups has shown that vibrotactile stimulation can be used for improved speech comprehension (Weisenberger & Percy, 1995, Cieśla et al., 2022, Fletcher et. al, 2019), localization of simple static sources (Gori et. al, 2014, Fletcher, Cunningham & Mills, 2020), and music perception (Branje et. al, 2010), both in TH individuals and in the hearing impaired/deaf subjects. We assume that this intuitive audio-tactile learning/substitution is due to the numerous similarities between these two senses, which is also the reason why the outcomes are so immediate (see *Introduction*; cf. long training regimes in. Bach-y-Rita et. al, 1969, Abboud et. al, 2014, Kupers & Ptito, 2011, Meijer, 1992).

Another parallel between the two sensory modalities can be drawn based on the subjective responses of our participants. When asked whether they were imagining/visualizing the tactile experience, 70% TH and 60% HI participants said “yes” (97% and 80% for the auditory stimuli, respectively). While it is a known phenomenon for auditory motion perception (Carlile & Leung, 2016), this result indicates a possible development of extrapersonal perception of tactile moving sources in some participants as well. This contributes to the general debate on whether objects rendered through SSDs can be perceived as existing directly in the external space, similar to the way natural senses operate (cf. distal attribution; Hartcher-O’Brien & Auvray, 2014, Maidenbaum et. al, 2014).

### Rehabilitation outlooks

Here we did not see significant improvement of the auditory motion localization (A2 vs A1) following a brief introduction of multisensory and tactile cues. However, we believe that dedicated longitudinal training protocols could help calibrate auditory perception (see similar suggestions in Gori et. al, 2014, Fletcher, Cunningham & Mills, 2020). This taking into account that a capability to adapt to experimentally altered binaural cues (e.g. through ear plugging) has been shown in a row of human and animal studies, and auditory localization capabilities can improve after acquiring experience with hearing devices (Shafiro et. al, 2015, Asp, Karltorp, & Berninger, 2022, Moore, 2002). An alternative multisensory training for spatial perception has also been suggested combining audio-visual inputs (Alzaher et. al, 2023, Valzolgher et. al, 2023, Aurelie 2022). The tactile sense handicap us that it can provide cues on the whole 360 azimuth without head or body motion, as opposed to vision. An audio-tactile intervention could potentially contribute to accelerating rehabilitation after CI or HA fitting, by refining the relationship between the various incoming auditory signals (e.g. electrical vs acoustic, or between two cochlear implants) as well as their relationship to spatial locations.

### Limitations of the study

In the current study, only one subgroup of the HI was homogeneous (10 users of BiCIs), while the rest of participants used various assistive device combinations: 3 were equipped with one CI and one HA, 4 had one CI and the other ear was effectively deaf, and 2 wore bilateral hearing aids. While we did not see any significant differences in performance between members of these subpopulations (see Figures 1-3,), more specific conclusions could arise by investigating them separately, while using more sizable groups. It would also be valuable to test the effects of variables, such as age at onset of hearing loss and its severity, duration of hearing loss and CI/HA use, on spatial localisation skills both through audition and touch. In addition, the intervention was rather short, i.e. only allowed for an acute assessment of spatial performance.

### Conclusions

Our findings support the role of both *nature* and *nurture* in the development of spatial understanding. For auditory spatial perception it seems that natural (or close to natural) cues are needed in order to develop proper spatial capabilities. Yet by using tactile means, we show that congenitally HI are nevertheless capable of representing 360° space. Furthermore, we show rapid integration of a new SSD-mediated sensory capability with existing sensory information. These findings have implications for neuroscience and are crucial towards further development of sensory rehabilitation for populations with hearing/sensory impairments.

## Material & Methods

The study was approved by the Reichman University Review Board (approval no 2022123) and conformed to the 2013 Helsinki Declaration. All participants signed an Informed Consent and were financially compensated for their participation. They were recruited through social media and with the help of previous participants.

### Participants

Nineteen (19) people with bilateral sensorineural hearing loss were included in the study (16 female, 3 male; age 26.5 +/− 8.09). Ten (10) were users of two cochlear implants (BiCI; 8 female, 2 male; age 27 +/− 6.4), three (3) were users of one cochlear implant and one hearing aid (biHA; bimodal stimulation), four (4) were users of a cochlear implant in one ear with the other ear deaf (>90dB HL for 0.25-8kHz range) and two (2) were users of two hearing aids. Hearing loss was of various etiology, in all participants diagnosed at birth (17/19) or within the first 5 years of life. None of the participants except for one (EG) had tinnitus. For the 3 users of a CI and a HA, in the ear with the HA the aided hearing thresholds for 250Hz-8kHz were 50-90 dB (EY), 30-70dB (YL), 40-80dB (TM); one user of two hearing aids had a mean binaural aided hearing threshold of 80-120 dB (RS), and one user of two bone-conduction devices had 20-70 dB (EG). All participants with hearing aids had a sloping hearing loss. Most users of cochlear implants got their first CI early in life, i.e. until the age of 3 and the second implant/a hearing aid before the age of 15 years. More specific details regarding the participants are depicted in Table 2. The used coded ID’s were not known to anyone outside the research group. The control group consisted of 29 individuals (22 female; 23.63+1.16; hereafter as TH). Randomly selected 22 people from the control group had a tonal audiometry test using an audiometer MAICO MA-51, with a staircase procedure of 5 dB up-10 dB down (in a soundbooth at the CANLAB laboratory of the Reichman University), and had normal hearing (<20dB for 250Hz-8kHz). The exclusion criteria for the study were neurological or psychiatric diseases, attention deficits, and tactile sensory issues.

**Table 2.**
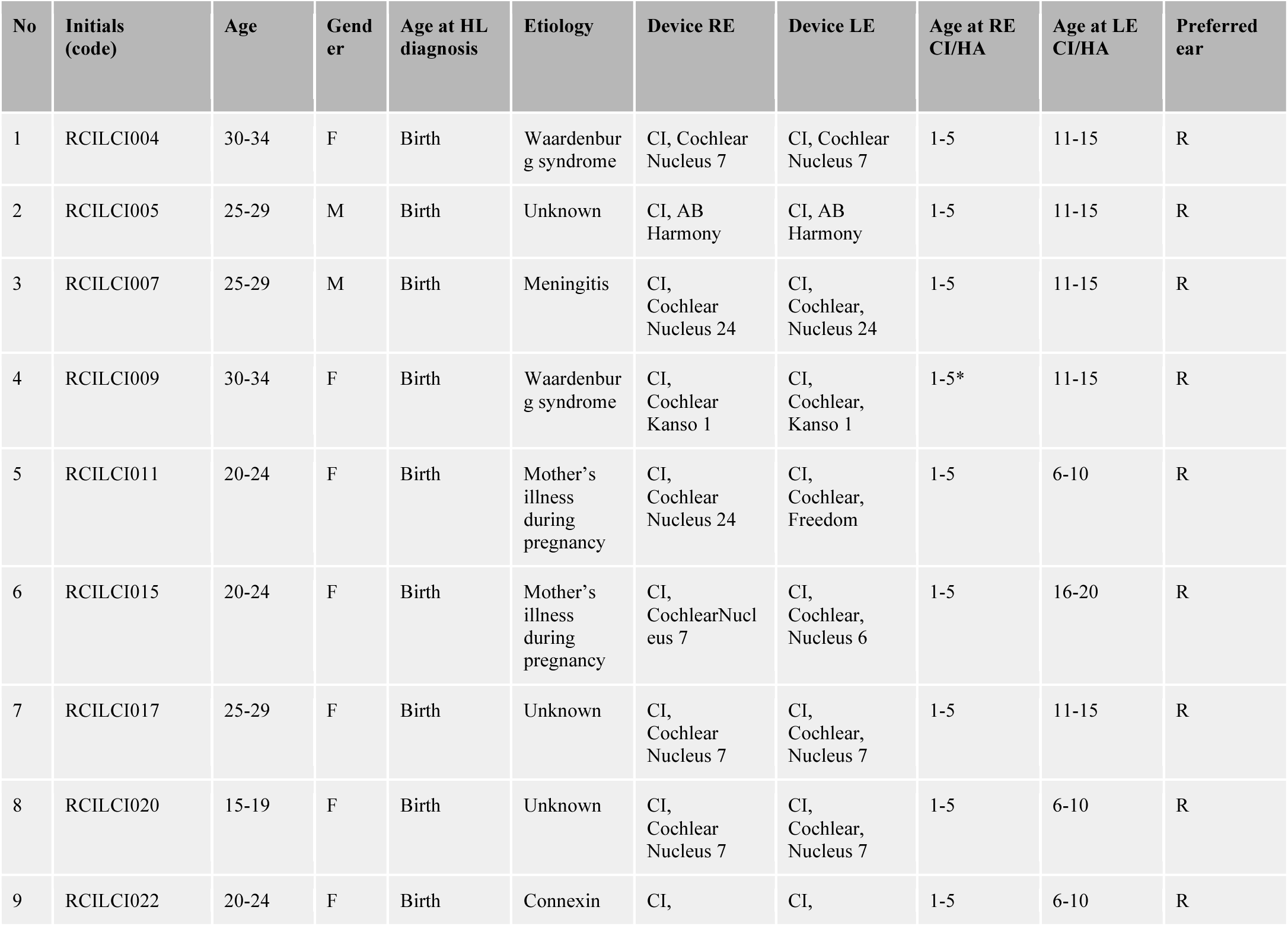
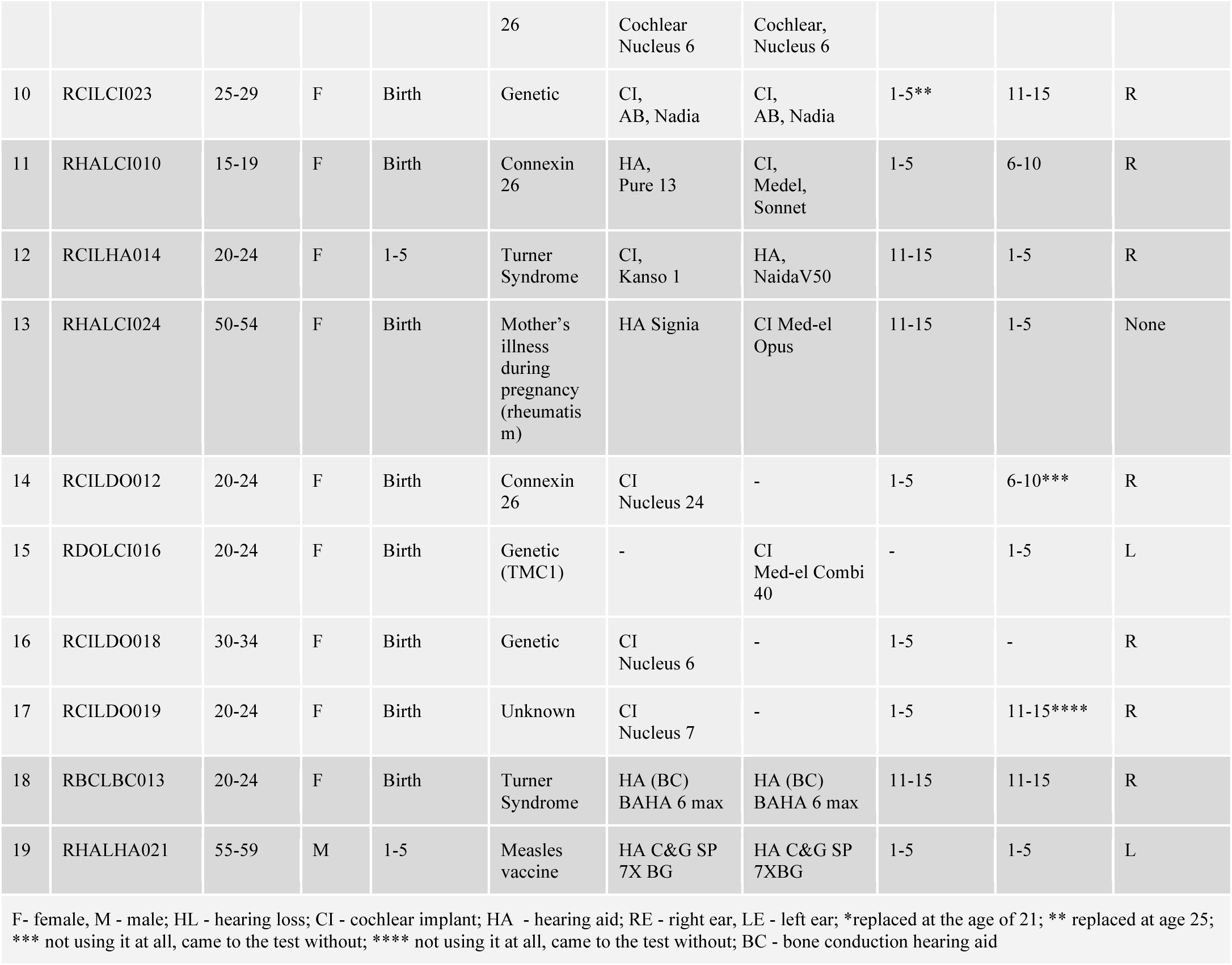
Details of the participants with hearing impairments.

### Experimental set-up

#### Apparatus

The experiment was conducted in a sound-treated cube-shaped room (4×4 meter), equipped with 97 loudspeakers (JBL Control 23-1L, powered by 13 Dante-enabled amplifiers, Crown Audio DCi 8|600DA) mounted on the walls and ceiling. The loudspeakers were organized in three horizontal rings, 24 loudspeakers each, at heights of 48cm, 148cm, 248cm from the ground and an azimuthal distance of 15° between adjacent speakers. The remaining twenty-five loudspeakers were set up on the ceiling in a 5×5 grid.

Auditory “moving” stimuli were decoded using an AllRad Ambisonics decoder (12th order) using the Spat5 library (version 5.2; Institut de Recherche et Coordination Acoustique/Musique, IRCAM) in the MaxMSP coding environment.

For the tactile stimulation, two in-house developed devices (VAS boxes) were used, each containing two piezoelectric plates to deliver vibrotactile stimulation on two fingertips (index and middle finger of both hands). The devices were sound-proofed and contained two silicone slits through which each finger was inserted and placed on top of a piezoelectric actuator (for further details see Cieśla et al., 2019, Cieśla et al., 2022).

One author of the manuscript (AS) developed an algorithm, TMA (Tactile Motion Algorithm), which decoded virtual positions of sounds in 360° space to four vibrotactile actuators of the VAS device. Each actuator represented a corner of the room (front-left, front-right, back-left, back-right). The algorithm performed vector-based level weighting among the four actuators, enabling smooth reproduction of the motion surrounding the participant (also possible due to the use of multifrequency tactile actuators). This in turn calculated the output level coefficient for each vibration actuator and set the content level accordingly (more details in: Snir & Ciesla, 2024). The content to the four tactile actuators was emitted as audio via the same Dante network as the Ambisonic sound environment. The algorithm and experimental paradigm were programmed in MaxMSP.

#### Preparations

Upon arrival at the Reichman University, each participant first signed an Informed Consent, and then was briefly interviewed regarding their demographic details and hearing status.

For the experimental part, the participant was seated in the center of the experimental room, with the head at the height of the center speaker ring and one tactile device on a small pedestal on the right and left side of the body (see Figures 1-2). The experimenter was seated in a separate control room and communicated with the participant using a talkback microphone (a microphone was placed at the ceiling of the experimental room). The experimenter could also monitor the participant using an overhead camera feed. In addition, all instructions were given to the participants in person face-to-face before each part of the experiment. The lights in the room were dimmed. Before any experiment started, it was also made sure that the participants could hear the experimental sounds inside the room, including the participants with hearing aids and cochlear implants (with their everyday device settings). All participants confirmed they could hear samples of sounds. The mobile phones of the hearing-impaired participants were placed in the control room, BlueTooth was disconnected.

#### Sound level measurements (dB levels)

Sound levels were measured in the experimental space using a MiniDSP UMIK-1 microphone (frequency response 8Hz-20kHz) placed at the participant’s head position. All measurements took place both with the tactile boxes ON and OFF. Specifically, the levels were measured for 20 randomly selected 4-second stimuli. The results were the following: a) tactile devices OFF: max. 65.3 +/− 2.2dB(z) vs b) tactile devices ON: max 64.6 +/−1.7dB(z).

### Experimental procedure

The stimuli for the experiment were constructed of a sawtooth wave at a frequency of 200Hz. Both audio and tactile stimuli were designed to induce perception of motion on a horizontal plane around the person. Motion could start in any of the eight positions around the participant, i.e. front, front-right, right, back-right, back, back-left, left, front-left; see Fig 2). The motion direction could be either clockwise or counter-clockwise. Each stimulus was 4 seconds in duration and could move at an azimuthal angle of 45°, 90°, 135°, 180°. There were four experimental conditions, in the following order for all individuals: Audio1 (A1), Tactile (T), Audio-Tactile (AT) and Audio2 (A2). Each condition had a separate test sheet containing 28 stimuli. Test sheets were pseudo-randomly created to equally represent the various orientations in space and directions of motion. Four catch trials included stimuli which were not moving at all. Responses were given verbally (e.g.“ The sound moved from front to right back”). Before the start of the experiment, each participant received an explanation of the localization task and was given a response sheet showing 8 possible responses (front, front-right, right, back-right, back, back-left, left, front-left). In addition, prior to the T condition a brief explanation of the tactile algorithm was given, and practiced with three example moving stimuli. For the T condition, CI users and HA users took their devices off, and the TH individuals wore headphones emitting white noise (due to the noise produced by the tactile device when no concurrent sounds are present). During the AT condition, the participants were not able to hear the noise of the tactile devices (see level measurements above).

### Data analysis

#### Task performance

The score for each condition in the localization task was calculated by combining the reported start-point, end-point and direction to arrive at the perceived midpoint of the motion trajectory, and comparing it to the actual stimulus midpoint. The distance in angle between the perceived and the actual stimulus midpoint was then converted to a score on a scale of 0-1 (perfect response: 0° error = 1; maximum error: 180° error = 0). Since localization was done on a circle and was calculated based on angle errors which to either direction would result as identical in scoring, a chance error was 0.5 (translating into 90° of error). The score was then compared between conditions (A1, AT, T, A2) for the TH group (N=29), the whole hearing-impaired group (N=19) and the bilateral cochlear implant users separately (N=10) (Wilcoxon Signed-Rank test). The midpoint scores for groups were also compared with one another, for each condition separately (Mann-Whitney tests). To assess front-back/back-front and right-left/left-right mistakes made during the localization tasks, the occurrences of such swaps with respect to the reported midpoints were calculated as percentage. The “front”, “back”, “left” and “right” subareas were defined as independent 90° quadrants (i.e. front = –45° ≥ mid-point ≥ 45°). In addition, the scores were presented as spatial orientation maps for each individual separately. These were arrived at by taking the scores of the midpoints for each of the sixteen possible angles (8 start points and 8 end points). These scores were then plotted using 16 point interpolation on a circle (the scipy.interpolate and matplotlib.pyplot libraries). Following each task (A1, AT, T, A2), participants were given a subjective questionnaire to assess certain aspects of their experience (Table 1).

#### Correlation Analysis

A Kendall r correlation analysis was applied to examine the relationship between the results in the experiment and age at fitting of both cochlear implants in bilateral CI users.

#### Additional questionnaires

##### Nijmegen Cochlear Implant Questionnaire (NCIQ) and Abbreviated Profile of Hearing Aid Benefit (APHAB)

To assess the subjective hearing status and the related quality of life (QoL) of the hearing impaired participants, one of two questionnaires were used depending on the used device. Participants with one or two cochlear implants (N=18) filled in the Nijmegen Cochlear Implant Questionnaire (NCIQ; Hinderink, Krabbe, & Van Den Broek, 2000). The original English version was translated to Hebrew by a bilingual English-Hebrew speaker. NCIQ is composed of 60 questions representing three hearing-related functional domains and (within them) 6 subdomains: 1) physical (basic sound perception, advanced sound perception, speech production), 2) psychological (self-esteem), 3) social (activity limitations, social interactions). Basic Sound Perception referred to hearing sounds like the phone, car, someone calling, wtc.; Advanced Sound Perception referred to situations, such as having a conversation, enjoying music, recognizing gender of the speaker, etc.; Speech Production referred to the ability of controlling one’s voice pitch and volume; Self-Esteem referred to accepting one’s own deficit, making new contacts, etc; Activity meant the amount of engagement at work, in hobbies, going out, etc.; Social Interaction referred to communications with different types of people and groups. The participants responded to the statements in the questionnaire as: never (1), sometimes (2), often (3), mostly (4), and always (5), or as no (1), poorly (2), moderate (3), adequate (4), and good (5) (last 5/60 items). An additional answer category was also offered, i.e. “not applicable (N/A)”. The score for a response to each item is: 1 = 0, 2 = 25, 3 = 50, 4 = 75, and 5 = 100. The total subdomain score is calculated by averaging the scores for 10 items per subdomain, with higher scores signifying better functioning. Participants who had either one or two hearing aids (N=5), filled in the Abbreviated Profile of Hearing Aid Benefit (APHAB, version A; Cox & Alexander, 1995). APHAB is an inventory for self-assessment, composed of 24 items concerning the experienced hearing-related handicap in everyday situations. The subjective benefit of the hearing aid is derived from comparing the results in the unaided vs aided condition. There are 4 subscales: Ease of Communication (EC), Reverberation (RV), Background Noise (BN), and Aversiveness (AV), with higher scores signifying more impaired functioning. The results are reported in the Supplementary Materials.

##### Colour Trial Test (CTT)

In order to exclude participants with sustained and divided attention deficits, the Colour Trial Test (CTT; d’Elia 1996) was performed by 15/19 participants with hearing deficits (3 males, 12 females, age: 29.13/– 11.55; 9 bilateral CI users, 1 bilateral HA user, 2 with 1 HA and 1 CI, 3 with 1 CI and deaf in the other ear) and 26 participants with typical hearing (7 males, 19 females, age: 23.37+/−1.4). Other participants couldn’t participate due to time constraints. The task is to connect circles following an ascending number sequence (1-25), while alternating between circles in yellow and circles in pink. The same numbers are presented both on the pink and on the yellow circles, therefore requiring that the participant ignores some of them. This attention test is as free as possible from the influences of language and cultural bias.

## Data Availability

All data produced in the present study are available upon reasonable request to the authors

## Acknowledgments

This research was supported by an ERC Consolidator Grant (773121 NovelExperiSense), a Horizon GuestXR (101017884) grant, and a Joy Ventures grant (to AA). We would like to thank Oran Goral for his support with the illustrations.

## Supplementary Materials

**Figure S1:**
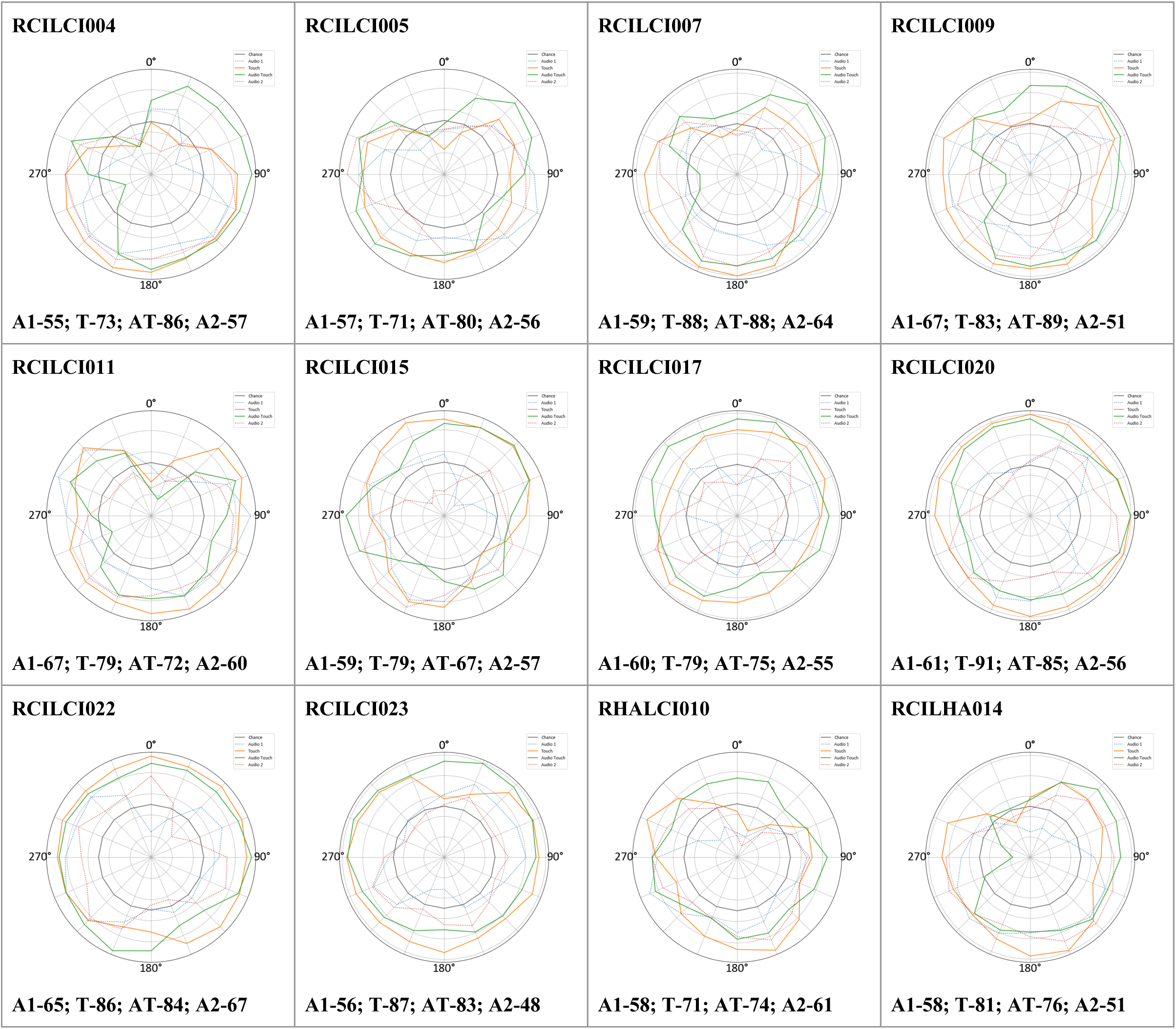
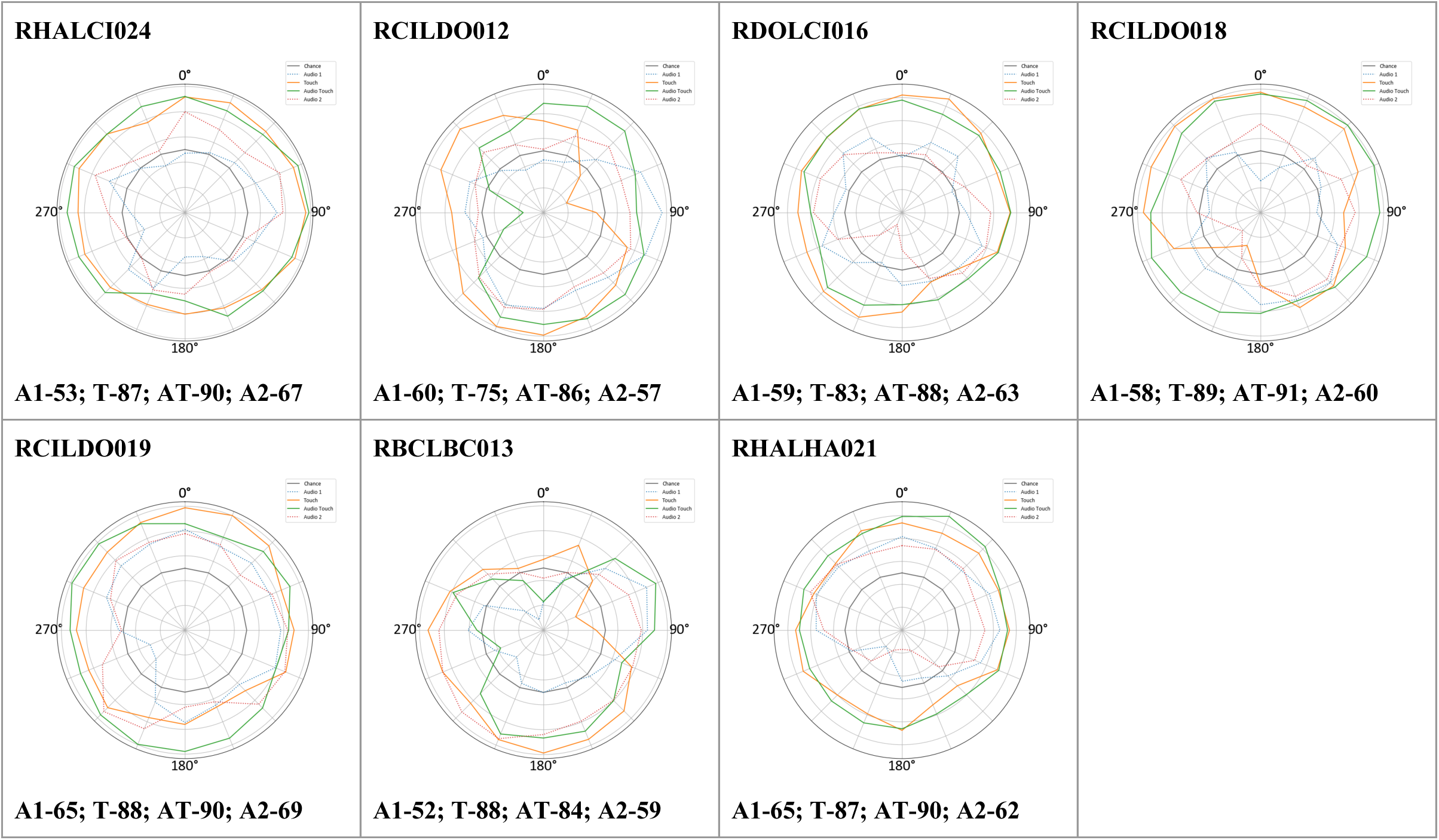
Individual spatial orientation maps in the CI/HA users; A1-first audio task, T – tactile task, AT – audio-tactile task, A2 – second audio task.

**Figure S2.**
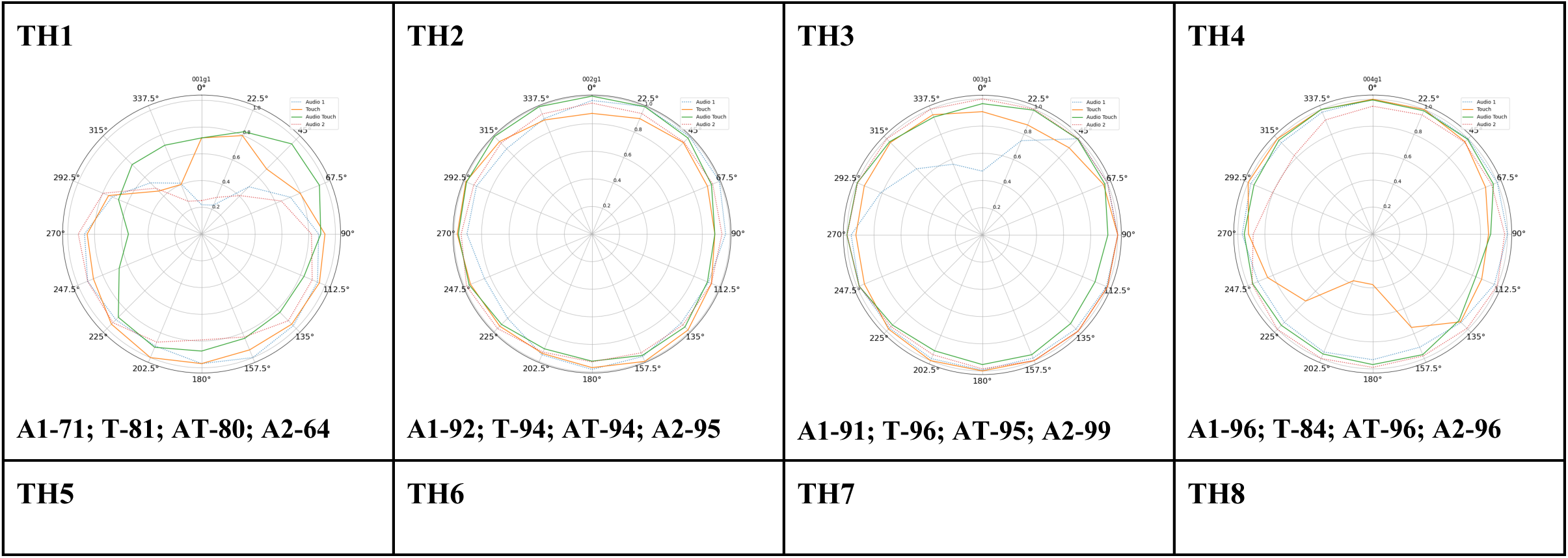
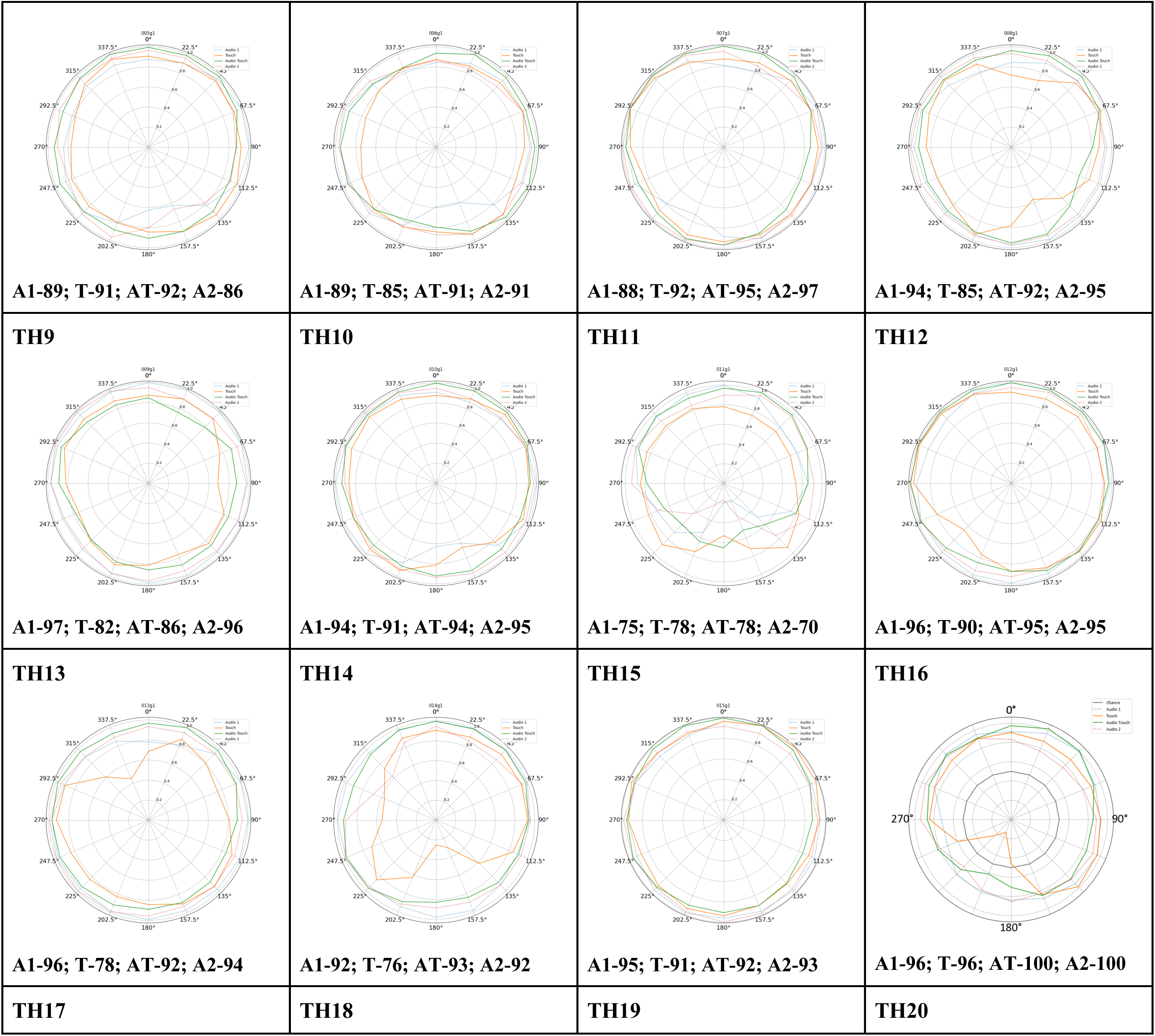
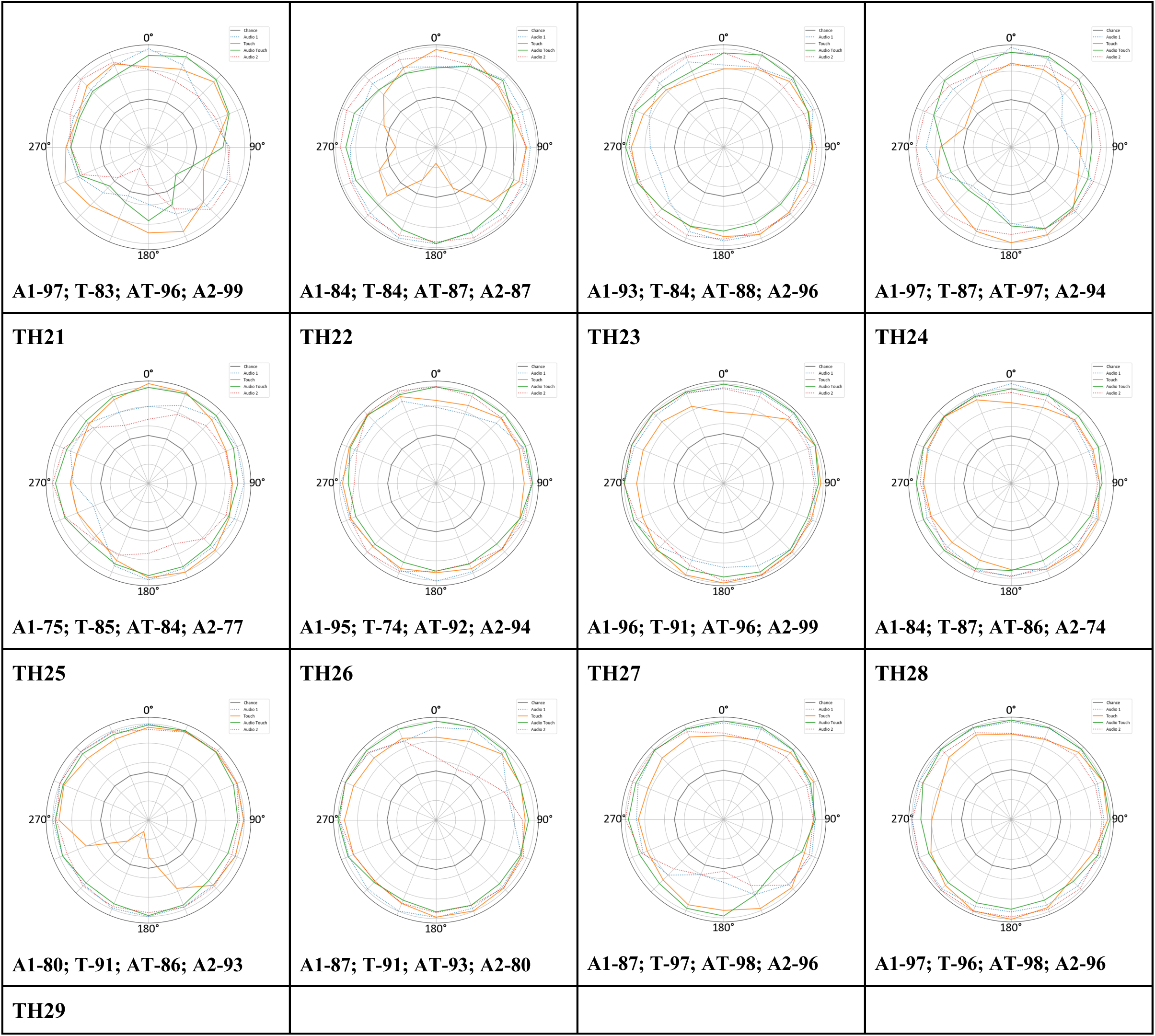
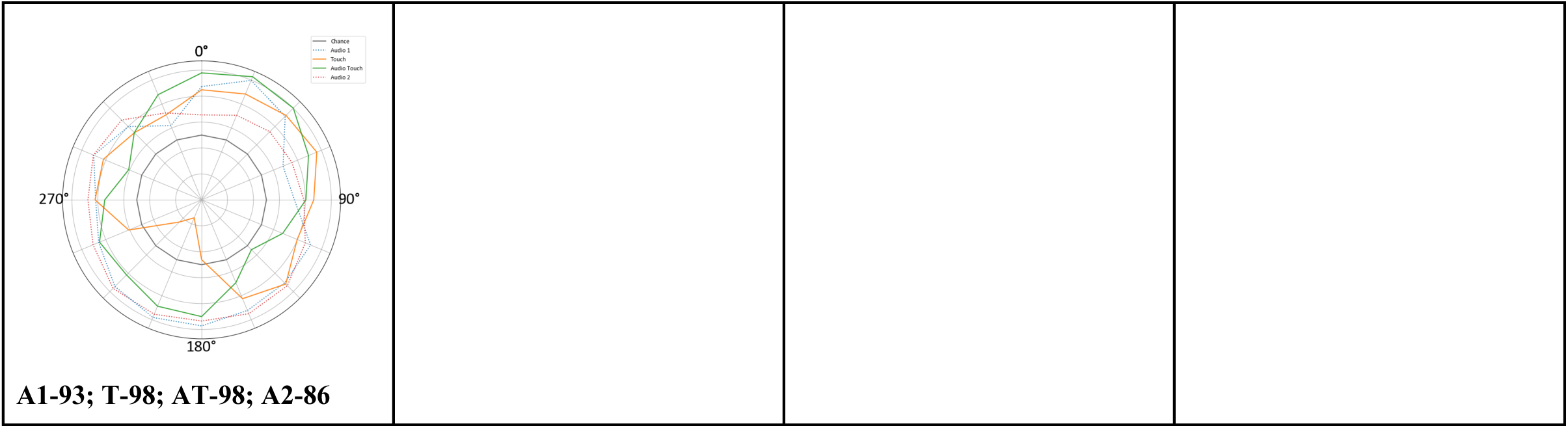
Individual spatial orientation maps in typically hearing individuals; A1 – first auditory condition, T – tactile condition, AT – audio-tactile condition, A2 – second auditory condition.

**Table S1.** Results of the APHAB questionnaire.

|  | HA<br>(EC) | HA<br>(RV) | HA<br>(BN) | HA<br>(AV) | noHA<br>(EC) | noHA<br>(RV) | noHA<br>(BN) | noHA<br>(AV) | Delta<br>(EC) | Delta<br>(RV) | Delta<br>(BN) | Delta<br>(AV) |
| --- | --- | --- | --- | --- | --- | --- | --- | --- | --- | --- | --- | --- |
| RHALCI010 | 71.38 | 44.28 | 61.11 | 52.53 | 83.67 | 48.48 | 79.46 | 7.07 | 12.29 | 4.21 | 18.35 | -45.45 |
| RCILHA014 | 40.07 | 56.73 | 75.42 | 25.76 | 100.00 | 77.44 | 95.96 | 1.01 | 59.93 | 20.71 | 20.54 | -24.75 |
| RHALCI024 | 33.84 | 50.51 | 50.51 | 56.57 | 56.57 | 48.48 | 79.46 | 1.01 | 22.73 | -2.02 | 28.96 | -55.56 |
| RBCLBC013 | 23.57 | 77.44 | 40.07 | 38.05 | 95.96 | 95.96 | 95.96 | 5.05 | 72.39 | 18.52 | 55.89 | -33.0 |
| RHALHA021 | 23.57 | 31.65 | 75.42 | 17.51 | 100 | 100 | 100 | 3.03 | 76.43 | 68.35 | 24.58 | -14.48 |

EC - Ease of Communication , RV - Reverberation, BN -Background Noise, AV - Aversiveness; a negative delta signifies no improvement with a HA/s on.

## References

1. Abboud, S., Hanassy, S., Levy-Tzedek, S., Maidenbaum, S., & Amedi, A. (2014). EyeMusic: Introducing a “visual” colorful experience for the blind using auditory sensory substitution. Restorative neurology and neuroscience, 32(2), 247–257.

2. Akeroyd, M. A. (2014). An overview of the major phenomena of the localization of sound sources by normal-hearing, hearing-impaired, and aided listeners. Trends in Hearing, 18, 2331216514560442.

3. Alzaher, M., Valzolgher, C., Verdelet, G., Pavani, F., Farnè, A., Barone, P., & Marx, M. (2023). Audiovisual training in virtual reality improves auditory spatial adaptation in unilateral hearing loss patients. Journal of Clinical Medicine, 12(6), 2357.

4. Alzaher, M., Valzolgher, C., Verdelet, G., Pavani, F., Farnè, A., Barone, P., & Marx, M. (2023). Audiovisual training in virtual reality improves auditory spatial adaptation in unilateral hearing loss patients. Journal of Clinical Medicine, 12(6), 2357.

5. Anderson, S. R., Jocewicz, R., Kan, A., Zhu, J., Tzeng, S., & Litovsky, R. Y. (2022). Sound source localization patterns and bilateral cochlear implants: Age at onset of deafness effects. PloS one, 17(2), e0263516.

6. Andersson, J. (2015). Horizontal sound localization in adults with unilateral or bilateral cochlear implants.

7. Arbel, R., Heimler, B., & Amedi, A. (2022). Face shape processing via visual-to-auditory sensory substitution activates regions within the face processing networks in the absence of visual experience. Frontiers in Neuroscience, 16, 921321.

8. Asp, F., Karltorp, E., & Berninger, E. (2022). Development of Sound Localization in Infants and Young Children with Cochlear Implants. Journal of Clinical Medicine, 11(22), 6758.

9. Bach-y-Rita, P., Collins, C. C., Saunders, F. A., White, B., & Scadden, L. (1969). Vision substitution by tactile image projection. Nature, 221(5184), 963–964.

10. Battal, C. (2018). Decoding auditory motion direction and location in hMT+/V5 and Planum Temporale of sighted and blind individuals.

11. Bavelier, D., Dye, M. W., & Hauser, P. C. (2006). Do deaf individuals see better?. Trends in cognitive sciences, 10(11), 512–518.

12. Beauchamp, M. S., Yasar, N. E., Frye, R. E., & Ro, T. (2008). Touch, sound and vision in human superior temporal sulcus. Neuroimage, 41(3), 1011–1020.

13. Best, V., Baumgartner, R., Lavandier, M., Majdak, P., & Kopčo, N. (2020). Sound externalization: A review of recent research. Trends in Hearing, 24, 2331216520948390.

14. Borg, E. (1997). Cutaneous senses for detection and localization of environmental sound sources: a review and tutorial. Scandinavian Audiology, 26(4), 195–206.

15. Branje, C., Maksimouski, M., Karam, M., Fels, D. I., & Russo, F. (2010, February). Vibrotactile display of music on the human back. In 2010 Third International Conference on Advances in Computer-Human Interactions (pp. 154-159). IEEE.

16. Bruns, P., & Röder, B. (2023). Development and experience-dependence of multisensory spatial processing. Trends in Cognitive Sciences.

17. Bruns, P., Li, L., Guerreiro, M. J. S., Shareef, I., Rajendran, S. S., Pitchaimuthu, K., Kekunnaya, R., & Röder, B. (2022). Audiovisual spatial recalibration but not integration is shaped by early sensory experience. iScience, 25(6), 104439. 10.1016/j.isci.2022.104439

18. Caetano, G., & Jousmäki, V. (2006). Evidence of vibrotactile input to human auditory cortex. Neuroimage, 29(1), 15–28.

19. Carlile, S., & Leung, J. (2016). The perception of auditory motion. Trends in hearing, 20, 2331216516644254.

20. Carlyon, R. P., & Goehring, T. (2021). Cochlear implant research and development in the twenty-first century: a critical update. Journal of the Association for Research in Otolaryngology, 22(5), 481–508.

21. Cieśla, K., Wolak, T., Lorens, A., Heimler, B., Skarżyński, H., & Amedi, A. (2019). Immediate improvement of speech-in-noise perception through multisensory stimulation via an auditory to tactile sensory substitution. Restorative neurology and neuroscience, 37(2), 155–166.

22. Cieśla, K., Wolak, T., Lorens, A., Mentzel, M., Skarżyński, H., & Amedi, A. (2022). Effects of training and using an audio-tactile sensory substitution device on speech-in-noise understanding. Scientific Reports, 12(1), 1–16.

23. Coudert, A., Gaveau, V., Gatel, J., Verdelet, G., Salemme, R., Farne, A., … & Truy, E. (2022). Spatial hearing difficulties in reaching space in bilateral cochlear implant children improve with head movements. Ear and Hearing, 43(1), 192–205.

24. Cox, R. M., & Alexander, G. C. (1995). The abbreviated profile of hearing aid benefit. Ear and hearing, 16(2), 176–186.

25. D’Elia, L., Satz, P., Uchiyama, C. L., & White, T. (1996). Color trails test. PAR.

26. Dorman, M. F., Loiselle, L. H., Cook, S. J., Yost, W. A., & Gifford, R. H. (2016). Sound source localization by normal-hearing listeners, hearing-impaired listeners and cochlear implant listeners. Audiology and Neurotology, 21(3), 127–131.

27. Dwyer, R. T., Chen, C., Hehrmann, P., Dwyer, N. C., & Gifford, R. H. (2021). Synchronized automatic gain control in bilateral cochlear implant recipients yields significant benefit in static and dynamic listening conditions. Trends in Hearing, 25, 23312165211014139.

28. Fischer, T., Schmid, C., Kompis, M., Mantokoudis, G., Caversaccio, M., & Wimmer, W. (2021). Pinna-imitating microphone directionality improves sound localization and discrimination in bilateral cochlear implant users. Ear and hearing, 42(1), 214–222.

29. Fletcher, M. D., Cunningham, R. O., & Mills, S. R. (2020). Electro-haptic enhancement of spatial hearing in cochlear implant users. Scientific Reports, 10(1), 1621.

30. Fletcher, M. D., Hadeedi, A., Goehring, T., & Mills, S. R. (2019). Electro-haptic enhancement of speech-in-noise performance in cochlear implant users. Scientific Reports, 9(1), 11428.

31. Gordon, K. A., Wong, D. D., & Papsin, B. C. (2013). Bilateral input protects the cortex from unilaterally-driven reorganization in children who are deaf. Brain, 136(5), 1609–1625.

32. Gori, M. (2015). Multisensory integration and calibration in children and adults with and without sensory and motor disabilities. Multisensory Research, 28(1-2), 71–99.

33. Gori, M., Campus, C., Signorini, S., Rivara, E., & Bremner, A. J. (2021). Multisensory spatial perception in visually impaired infants. Current Biology, 31(22), 5093–5101.

34. Gori, M., Vercillo, T., Sandini, G., & Burr, D. (2014). Tactile feedback improves auditory spatial localization. Frontiers in Psychology, 5, 109563.

35. Hartcher-O’Brien, J., & Auvray, M. (2014). The process of distal attribution illuminated through studies of sensory substitution. Multisensory Research, 27(5-6), 421–441.

36. Heimler, B., & Amedi, A. (2020). Are critical periods reversible in the adult brain? Insights on cortical specializations based on sensory deprivation studies. Neuroscience & Biobehavioral Reviews, 116, 494–507.

37. Heimler, B., Striem-Amit, E., & Amedi, A. (2015). Origins of task-specific sensory-independent organization in the visual and auditory brain: neuroscience evidence, open questions and clinical implications. Current opinion in neurobiology, 35, 169–177.

38. Hinderink, J. B., Krabbe, P. F., & Van Den Broek, P. (2000). Development and application of a health-related quality-of-life instrument for adults with cochlear implants: the Nijmegen cochlear implant questionnaire. Otolaryngology–Head and Neck Surgery, 123(6), 756–765.

39. Holmes, N. P. (2007). The law of inverse effectiveness in neurons and behaviour: multisensory integration versus normal variability. Neuropsychologia, 45(14), 3340–3345.

40. Hubel, D. H., & Wiesel, T. N. (1962). Receptive fields, binocular interaction and functional architecture in the cat’s visual cortex. The Journal of physiology, 160(1), 106.

41. Johnson, J. A., Xu, J., & Cox, R. M. (2017). Impact of hearing aid technology on outcomes in daily life III: Localization. Ear and Hearing, 38(6), 746–759.

42. Kalmijn, A. J. (1989). Functional evolution of lateral line and inner ear sensory systems. In The mechanosensory lateral line: neurobiology and evolution (pp. 187–215). New York, NY: Springer New York.

43. Kamke, M. R., Van Luyn, J., Constantinescu, G., & Harris, J. (2014). Contingent capture of involuntary visual spatial attention does not differ between normally hearing children and proficient cochlear implant users. Restorative Neurology and Neuroscience, 32(6), 799–811.

44. Kan, A., & Litovsky, R. Y. (2015). Binaural hearing with electrical stimulation. Hearing Research, 322, 127–137.

45. Kayser, C., Petkov, C. I., Augath, M., & Logothetis, N. K. (2005). Integration of touch and sound in auditory cortex. Neuron, 48(2), 373–384.

46. King, A. (2004). Development of multisensory spatial integration. Crossmodal space and crossmodal attention, 1–24.

47. Kral, A., & Sato, M. (2020). Nature and nurture in hearing: critical periods for therapy of deafness. Acoustical Science and Technology, 41(1), 54–58.

48. Kupers, R., & Ptito, M. (2011). Cross-modal brain plasticity in congenital blindness: lessons from the tongue display unit. i-Perception, 2(8), 748–748.

49. Laback, B., Egger, K., & Majdak, P. (2015). Perception and coding of interaural time differences with bilateral cochlear implants. Hearing Research, 322, 138–150.

50. Landry, S. P., Guillemot, J. P., & Champoux, F. (2013). Temporary deafness can impair multisensory integration: a study of cochlear-implant users. Psychological science, 24(7), 1260–1268.

51. Landry, S. P., Guillemot, J. P., & Champoux, F. (2014). Audiotactile interaction can change over time in cochlear implant users. Frontiers in Human Neuroscience, 8, 316.

52. Li Hegner, Y., Lee, Y., Grodd, W., & Braun, C. (2010). Comparing tactile pattern and vibrotactile frequency discrimination: a human FMRI study. Journal of neurophysiology, 103(6), 3115–3122.

53. Litovsky, R. (2015). Development of the auditory system. Handbook of clinical neurology, 129, 55–72.

54. Lu, L., Zhang, X., & Gao, X. (2019). Non-implantable Artificial Hearing Technology. Hearing Loss: Mechanisms, Prevention and Cure, 145–163.

55. Ludwig, A. A., Meuret, S., Battmer, R. D., Schönwiesner, M., Fuchs, M., & Ernst, A. (2021). Sound localization in single-sided deaf participants provided with a cochlear implant. Frontiers in psychology, 12, 753339.

56. Lundbeck, M., Grimm, G., Hohmann, V., Laugesen, S., & Neher, T. (2017). Sensitivity to angular and radial source movements as a function of acoustic complexity in normal and impaired hearing. Trends in hearing, 21, 2331216517717152.

57. Maidenbaum, S., Hanassy, S., Abboud, S., Buchs, G., Chebat, D. R., Levy-Tzedek, S., & Amedi, A. (2014). The “EyeCane”, a new electronic travel aid for the blind: Technology, behavior & swift learning. Restorative neurology and neuroscience, 32(6), 813–824.

58. McLachlan, G., Majdak, P., Reijniers, J., & Peremans, H. (2021). Towards modelling active sound localisation based on Bayesian inference in a static environment. Acta Acustica, 5, 45.

59. Meijer, P. B. (1992). An experimental system for auditory image representations. IEEE transactions on biomedical engineering, 39(2), 112–121.

60. Meredith, M. A., & Stein, B. E. (1983). Interactions among converging sensory inputs in the superior colliculus. Science, 221(4608), 389–391.

61. Middlebrooks, J. C. (2015). Sound localization. Handbook of clinical neurology, 129, 99–116.

62. Moore, D. R. (2002). Auditory development and the role of experience. British Medical Bulletin, 63(1), 171–181.

63. Moua, K., Kan, A., Jones, H. G., Misurelli, S. M., & Litovsky, R. Y. (2019). Auditory motion tracking ability of adults with normal hearing and with bilateral cochlear implants. The Journal of the Acoustical Society of America, 145(4), 2498–2511.

64. Mueller, M. F., Meisenbacher, K., Lai, W. K., & Dillier, N. (2014). Sound localization with bilateral cochlear implants in noise: How much do head movements contribute to localization?. Cochlear implants international, 15(1), 36–42.

65. Nava, E., Bottari, D., Villwock, A., Fengler, I., Büchner, A., Lenarz, T., & Röder, B. (2014). Audio-tactile integration in congenitally and late deaf cochlear implant users. PLoS One, 9(6), e99606.

66. Nisha, K. V., Uppunda, A. K., & Kumar, R. T. (2023). Spatial rehabilitation using virtual auditory space training paradigm in individuals with sensorineural hearing impairment. Frontiers in Neuroscience, 16, 1080398.

67. O’Connell-Rodwell, C. E. (2007). Keeping an “ear” to the ground: seismic communication in elephants. Physiology.

68. Pastore, M. T., Natale, S. J., Yost, W. A., & Dorman, M. F. (2018). Head movements allow listeners bilaterally implanted with cochlear implants to resolve front-back confusions. Ear and hearing, 39(6), 1224–1231.

69. Pavani, F., Venturini, M., Baruffaldi, F., Artesini, L., Bonfioli, F., Frau, G. N., & van Zoest, W. (2017). Spatial and non-spatial multisensory cueing in unilateral cochlear implant users. Hearing research, 344, 24–37.

70. Rasmussen, K. M. B., West, N. C., Bille, M., Sandvej, M. G., & Cayé-Thomasen, P. (2022). Cochlear implantation improves both speech perception and patient-reported outcomes: a prospective follow-up study of treatment benefits among adult cochlear implant recipients. Journal of Clinical Medicine, 11(8), 2257.

71. Rezk, M., Cattoir, S., Battal, C., Occelli, V., Mattioni, S., & Collignon, O. (2020). Shared representation of visual and auditory motion directions in the human middle-temporal cortex. Current Biology, 30(12), 2289–2299.

72. Ro, T., Ellmore, T. M., & Beauchamp, M. S. (2013). A neural link between feeling and hearing. Cerebral cortex, 23(7), 1724–1730.

73. Santos, N. P. D., Couto, M. I. V., & Martinho-Carvalho, A. C. (2017, December). Nijmegen Cochlear Implant Questionnaire (NCIQ): translation, cultural adaptation, and application in adults with cochlear implants. In Codas (Vol. 29, p. e20170007). Sociedade Brasileira de Fonoaudiologia.

74. Schäfer, E., Vedoveli, A. E., Righetti, G., Gamerdinger, P., Knipper, M., Tropitzsch, A., … & Li Hegner, Y. (2021). Activities of the right temporo-parieto-occipital junction reflect spatial hearing ability in cochlear implant users. Frontiers in Neuroscience, 15, 613101.

75. Schnupp, J., Nelken, I., & King, A. (2011). Auditory neuroscience: Making sense of sound. MIT press.

76. Shafiro, V., Sheft, S., Kuvadia, S., & Gygi, B. (2015). Environmental sound training in cochlear implant users. Journal of speech, language, and hearing research, 58(2), 509–519.

77. Snir, A., Ciesla, K., Vekslar, R., Ozdemir, G., & Amedi, A. (2024). Localizing 3D motion through fingertips: following in the footsteps of elephants [in press IScience]

78. Spence, C. (2010). Crossmodal spatial attention. Annals of the New York Academy of Sciences, 1191(1), 182–200.

79. Spence, C., & Di Stefano, N. (2024). What, if anything, can be considered an amodal sensory dimension?. Psychonomic Bulletin & Review, 1–19.

80. Tranchant, P., Shiell, M. M., Giordano, M., Nadeau, A., Peretz, I., & Zatorre, R. J. (2017). Feeling the beat: Bouncing synchronization to vibrotactile music in hearing and early deaf people. Frontiers in neuroscience, 11, 281459.

81. v Békésy, G. (1957). Sensations on the skin similar to directional hearing, beats, and harmonics of the ear. The Journal of the Acoustical Society of America, 29(4), 489–501.

82. Valzolgher, C., Gatel, J., Bouzaid, S., Grenouillet, S., Todeschini, M., Verdelet, G., … & Pavani, F. (2023). Reaching to sounds improves spatial hearing in bilateral cochlear implant users. Ear and Hearing, 44(1), 189–198.

83. Warnecke, M., Peng, Z. E., & Litovsky, R. Y. (2020). The impact of temporal fine structure and signal envelope on auditory motion perception. PLoS One, 15(8), e0238125.

84. Weisenberger, J. M., & Percy, M. E. (1995). The transmission of phoneme-level information by multichannel tactile speech perception aids. Ear and hearing, 16(4), 392–406.

85. Yost, W. A., Zhong, X., & Najam, A. (2015). Judging sound rotation when listeners and sounds rotate: Sound source localization is a multisystem process. The Journal of the Acoustical Society of America, 138(5), 3293–3310.

86. Yu, H. V., Tao, L., Llamas, J., Wang, X., Nguyen, J. D., Trecek, T., & Segil, N. (2021). POU4F3 pioneer activity enables ATOH1 to drive diverse mechanoreceptor differentiation through a feed-forward epigenetic mechanism. Proceedings of the National Academy of Sciences, 118(29), e2105137118.

87. Zheng, Y., Swanson, J., Koehnke, J., & Guan, J. (2022). Sound localization of listeners with normal hearing, impaired hearing, hearing aids, bone-anchored hearing instruments, and cochlear implants: a review. American journal of audiology, 31(3), 819–834.

